# Peripheral Treg-monocyte immune signatures relate to neurodegeneration and prognosis in patients with primary tauopathies

**DOI:** 10.64898/2026.03.17.26348492

**Authors:** Kei Onn Lai, Julia Goddard, Harry Crook, Roosmarijn Frohn, Stacey L Kigar, Natalia Savinykh Yarkoni, Peter Swann, Robert Durcan, Julie Wiggins, Win Li, Hugo Paula, Timothy Rittman, Amanda Heslegrave, James B Rowe, Matthias Brendel, Henrik Zetterberg, Josef Priller, John T O’Brien, Maura Malpetti

## Abstract

**Background:** Neuroinflammation is a common hallmark of primary tauopathies, and is associated with worse clinical outcomes over time. However, accurate prognosis in these disorders remains challenging, and current fluid biomarkers provide limited insight into the contribution of peripheral immune cells to PSP/CBS pathogenesis. Our study aims to characterise blood-based immune cell profiles in patients with progressive supranuclear palsy (PSP) and corticobasal syndrome (CBS), and test their associations with neurodegeneration and clinical outcomes.

**Methods:** Peripheral blood immune cells from fresh whole blood were characterized with high-dimensional mass cytometry (29 markers) in n=60 people with PSP/CBS and n=21 age- and sex-matched controls. Cell type abundance was defined as the ratio of counts for each gated population divided by total live cells. Hierarchical clustering of cell types and principal component analysis were used to derive data-driven immune clusters. Correlation network analysis and diffusion-based network propagation integrated cell counts with plasma inflammation markers to prioritise mediators of intercellular signalling. Associations between immunological markers, plasma concentrations of neurofilament light chain (NfL), cognition, and survival were assessed using regression and Cox proportional hazards models.

**Results:** Patients with PSP/CBS showed a global increase in covariance among immune cell populations, indicating heightened coordination within the peripheral immune network. A monocyte-driven cluster (Cluster 1) showed higher scores in PSP/CBS, reflecting impaired phenotypic transition from classical to nonclassical monocytes, and was associated with higher NfL levels, poorer cognitive performance, and worse prognosis. In contrast, a Treg-driven cluster (Cluster 2) showed lower scores in PSP/CBS, and was associated with better cognition and longer survival. Integrated multimodal networks identified a small set of immune-regulatory molecules and cytokines mediating crosstalk between Treg/Th17-like cells and monocytic populations, supporting a dysregulated Treg–monocyte axis in PSP/CBS.

**Conclusions:** We identified peripheral blood-based immunophenotypic profiles of individuals with PSP/CBS that are associated with neurodegeneration, cognitive decline, and survival. Dysregulated monocyte maturation and reduced Treg-related immune configurations are enriched in patients with worse outcomes, suggesting that specific peripheral immune cell subsets may serve as fluid biomarkers and potential immunotherapy targets in primary tauopathies.

## Background

Progressive supranuclear palsy (PSP) and Corticobasal syndrome (CBS) are clinical syndromes often underpinned by primary tauopathies – including PSP pathology and corticobasal degeneration (CBD) - whereby four-repeat (4R) tau aggregates are found in intracellularly in neurons and astroglia [1, 2]. These conditions are relentlessly progressive, with death occurring on average ∼5-6 years after symptom onset [1]. Primary tauopathies, similar to other neurodegenerative diseases like Alzheimer’s disease (AD), are characterized by immune activation as an early driver of pathogenesis, evidenced by activated microglia and inflammatory signatures that colocalize with neurodegeneration in postmortem studies [3–7] and animal models [8–11]. Early associations between neuroinflammation and tauopathies have been confirmed by large scale GWAS cohort studies where several glial activation genes, such as *LRRK2, SLCO1A2* and *RUNX2*, were identified as PSP risk genes [12–14].

*In vivo* PET imaging in patients with PSP and CBS have described strong associations between neuroinflammation in core pathological regions, clinical severity and faster decline over time [15–19]. Beyond the brain, recent evidence from fluid-based proteomics analyses have identified immune signatures in cerebrospinal fluid [20–24] and blood of patients with PSP and CBS [19, 25–27], including elevated levels of pro-inflammatory cytokines that are associated with neuroinflammation and shorter survival (e.g. TNF-α, IL-6,IL-17A) [25, 26, 28, 29]. Although proteomics analyses are a powerful tool to identify inflammatory signatures, they are often cell type agnostic and cannot provide a full molecular framework on disease pathogenesis. Complementing these molecular approaches, cell type profiling has revealed distinct peripheral and central immune cell alterations in neurodegenerative diseases [30–33]. For example, tau spread was found to be modulated by the interaction of CD8+ T cells with microglia in preclinical mouse models [32]. Besides the brain parenchyma, growing evidence has shown that peripheral immune cells are clinically relevant to neurodegenerative diseases. For example, peripheral immune cells profiling has been used in stratification of Parkinson’s Disease [33]. For PSP/CBS, reduced abundance of nonclassical monocytes paired with increases in classical monocytes in the peripheral blood was associated with shorter survival in patients [30]. Similarly, high neutrophil-to-lymphocyte ratio in blood is associated with worse disease severity in patients with PSP [34]. Immunophenotyping studies of peripheral immune cells remain scarce in patients with PSP and CBS, and are restricted to low-dimension marker profiles or few cell types [30, 34–38]. Nevertheless, these previous studies suggest an active contribution of peripheral immune cells on disease pathogenesis and clinical progression in PSP/CBS.

Defining immunophenotypic profiles in patients with primary tauopathies reflect a critical step to identify targetable pathways, to validate tools to stratify patients, and to empower new therapeutic strategies and trials. In this context, our study aims to identify clinically relevant blood immune cell signatures in PSP and CBS using high-dimensional immune cell profiling linked to disease severity and clinical progression. We hypothesise that specific peripheral immune cell configurations may constitute prognostic proxies in PSP and CBS, capturing disease-relevant biology that complements imaging and other fluid biomarkers and ultimately informing patient selection, stratification and outcome measures in future immunomodulatory trials.

## Materials and Methods

### Participants

We recruited 60 patients with a clinical diagnosis of PSP (n=49) [39] or CBS (n=11) [40], as part of specialist clinics for cognitive and movement disorders at the Cambridge University Hospitals National Health Service Trust, and 21 age- and sex-matched healthy volunteers.

Exclusion criteria for recruitment included diagnoses of autoimmune diseases, cancer, major concurrent psychiatric illness or other severe physical illness; cancer-related therapy within the past 4 years; infections, surgery, operations or vaccinations in the past 4 weeks; or a history of other significant neurological illness.

All participants underwent phlebotomy, clinical and cognitive assessment. Both EDTA and sodium heparin blood collection tubes were used to collect blood samples obtained by venepuncture. For cognitive assessments, participants performed the revised Addenbrooke’s Cognitive Examination (ACER, 0-100 scale) [41], which tests five cognitive subdomains, including Fluency (Maximum 14 points), Attention and Orientation (Maximum 18 points), Memory (Maximum 26 points), Language (Maximum 26 points) and Visuospatial (Maximum 16 points), with lower scores indicating worse cognitive performance. To evaluate clinical severity in patients, the PSP Rating Scale (PSP-RS) was performed, testing 6 domains (history/daily activities, mentation, bulbar motor, ocular motor, limb motor and gait areas). Higher PSP-RS scores indicate worse disease severity, with a maximum total score of 100.

Participants with mental capacity gave their written informed consent to take part in the study according to the Declaration of Helsinki. For those who lacked capacity, their participation followed the consultee process in accordance with UK law. The research protocols were approved by the National Research Ethics Service’s East of England Cambridge Central Committee.

### Plasma sample processing and NULISAseq analyses

Participant’s blood samples in EDTA tubes were centrifuged at 2000rpm, for 10min at 4°C, to isolate plasma, which were then kept in 250 µLs aliquots at −70 °C until subsequent analyses with the NULISAseq [42] assays. The NULISA CNS panel enables detection of 120 CNS fluid biomarkers including neurodegenerative biomarkers, including phosphorylated tau forms (e.g. p-tau217) and neurofilament light chain (NfL). In addition, the NULISAseq Inflammation Panel enables simultaneous analyses of 250 proteins associated with inflammatory and immune response processes (e.g. interleukins, chemokines, complement). Sample and data analysis was performed according to kit manufacturer protocols, including Log2 Transformation of the data and NULISA Protein Quantification (NPQ) on the logarithmic scale (Alamar Biosciences, Fremont, CA). See our previous study [25] for more details.

### Peripheral blood mononuclear cell (PBMC) immunophenotyping by mass cytometry

On the same day of blood collection, 500 μL aliquot of sodium-heparin whole blood was prepared for mass cytometry following the Maxpar Direct Immune Profiling (DIP) Assay manufacturer’s guidelines (Cat# 201334, Standard BioTools). Briefly, samples were treated for 20 mins with 0.15 kilo-units of sodium-heparin salt (Cat#: H3149-10KU, Sigma Aldrich) diluted in Maxpar PBS (Cat#: S00125, Standard BioTools). 255μL of this heparin-blocked blood was then added directly to a Maxpar DIP Assay tube, pre-manufactured with lyophilized antibody cocktail. Antibody incubation proceeded for 30 mins, then 250 μL of Cal-Lyse™ solution (Cat#: GAS-010S100, Thermo Fisher) was added for 10 mins to lyse red blood cells. Next, samples were diluted with 3mL of MilliQ water, incubated for a further 10 mins at room temperature until the cell suspension became translucent, and were centrifuged at 300g for 5 mins. After discarding the supernatant, the sample was resuspended in 3mL of Maxpar CSB (Cat#: 201068, Standard BioTools) and centrifuged at 300g for 5 mins (RT) for a total of three washes. The sample were then fixed in 1.6% formaldehyde solution (16% formaldehyde, Cat#: 28908, Pierce; diluted 1:10 in Maxpar PBS) for 10 mins, spun at 800g for 5 mins, and decanted. The samples were then resuspended in 1 mL of freshly prepared freezing media consisting of 10% DMSO (Cat# 10103484, Fisher Scientific Ltd) in AB serum (Cat # H4522-100ml, Merck Life Science Ltd), transferred to cryovials, placed into pre-cooled CoolCell, and immediately frozen at -70°C.

All pre-processed samples were grouped into batches of 6-7 samples and analysed within ∼6 weeks post collection. On the day before acquisition, samples were thawed in a 37°C water bath. Maxpar CSB (2mL) was added to the thawed samples, gently vortexed, and the samples were spun at 800g for 5 mins. After discarding the supernatant, the samples were resuspended in 1 mL of 125nM Cell-ID Intercalator-Ir (Cat# 201240, Standard BioTools) in Maxpar Fix & Perm buffer (Cat#: S00092, Standard BioTools) and left for staining overnight in the fridge at 4°C. The next day, samples were centrifugated at 800g for 5 mins. After discarding the supernatant, the samples were resuspended in 2 mL of Maxpar CSB buffer and centrifuged at 800g for 5 mins. Maxpar CSB wash step were repeated twice, followed by two more washes with 2ml Maxpar CAS (Cat#: 201240, Standard BioTools) at 800g for 5 mins. Before the last centrifugation step, a 10 μL aliquot was taken for cell counting. Samples were then taken for data acquisition using a Helios time of flight (TOF) mass cytometer. Unless otherwise noted, all steps were performed at room temperature.

From the raw fcs mass cytometry files, bead normalization was performed to extract the stable signals across time through the instrument channels using *CATALYST* R package. Identification and extraction of the single live cells data were performed using the cytofQC support vector machine model. Gating was performed as guided by *Maxpar Direct Immune Profiling application protocol* [43] using flowWorkspace and *CytoML* R packages. Gating (*Supplementary Figures 1-2, gating hierarchy with 29 cell surface markers*) was performed in a blinded manner to groups of the individuals.

### Statistical analyses

Analyses were performed using R (version 4.4.1). For descriptive statistics, chi-square and two-sided permutational tests were used to compare variables between the PSP/CBS and control groups. Analysis of variance on brain-derived p-tau-217 plasma levels was applied to compare concentrations between patients with PSP, patients with CBS and controls, to test for AD (co-)pathology. In fact, although most patients with CBS present with primary tauopathies, including CBD and PSP, a portion of cases can be underpinned by AD pathology(∼30-40%) [44, 45]. Blood-derived immunophenotyping cell quantification values were extracted from 60 bivariate gates (*Supplementary Figures 1-2 for gating hierarchy*). Throughout the recruitment, we excluded participants diagnosed with inflammatory conditions. We also compared levels of plasma C-reactive proteins (CRP) of the recruited individuals, to evaluate possible confounding signal of pre-existing systemic inflammation for between groups comparison.

Across all analysis steps, statistical significance was defined as p values <0.05, and p values were corrected with False Discovery Rate (FDR) correction when multiple comparisons occurred.

#### Differential abundance of cell types and cell type clusters

Immune cell analyses included 27 cell types from high dimensional mass cytometry data. Generalized negative binomial models were implemented to determine if individual cell type abundance differ across groups (patients vs controls). Age, sex and batches of mass cytometry run (“Helios batch”) were added as covariates. Log transformed total live cells were added as offset to account for sampling differences. Group 𝛽 coefficient represents relative differences in cell type abundance.

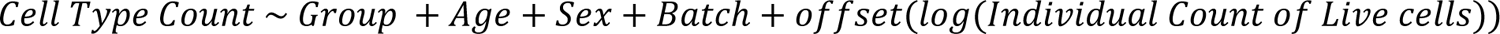

To investigate immune cell coordination dynamics and covariance, we applied clustering analyses on the abundance of the cell types. Firstly, pairwise complete Pearson correlations were applied across all cell populations. Positive correlations were kept and the related dissimilarity index was calculated (1-correlation) for each correlation pair. Then, complete linkage clustering based on the dissimilarity matrix was performed. The number of clusters to retain (“k”) was obtained based on silhouette analyses, considering the most parsimonious solution that also aligned with the classification of major immune cell types. To extract the summary of cell abundance represented in each cluster, principal component analyses were performed within clusters. For each cluster, we then considered the first Principal Component score as an abundance metric of each participant. Two-sided permutational test (10000 iterations, using *MKinfer* R package), were used to evaluate group differences in each cluster’s component scores.

#### Correlational network analyses of immune cell type abundance

Partial Pearsons correlation matrices across cell sub-types (abundance) controlling for age, sex and Helios batch were calculated. The matrices were converted into Circos plots, where only significant edge links were represented (r >0.3, *FDR-corrected p <0.05*). For network analyses, undirected weighted graphs were built from the significant positive correlations. Weighted degrees were calculated based on the sum of positive correlation strengths for single node (cell type). Betweenness centralities were used to define how often a node lies on shortest paths between others within the undirected graph. Weights of betweenness centralities were based on correlation strengths. All network metrics were based on *igraph* R package.

#### Differential expression of cell surface markers

Geometric median mass isotope intensities were used as a proxy for protein marker expression in of cell populations of interest using the *getStats* function from *flowWorkspace* R package. Intensities were then corrected for batch effects using the *removebatchEffect* function from *limma* R package. Two-sided permutational t-test (n=10000 iterations, *MKinfer* R package) was performed to evaluate marker expression differences between patients and controls. Radar plots were created to illustrate multivariate marker expression patterns and enable group comparisons for specific cell types. For each cell type, standardized median expression per marker was plotted. To ensure comparability of expression within the cell type, radial shell axes were based on the global minimum and maximum expression across all the markers. For each cell type, only positive markers as defined by the Maxpar gating strategy [43], are included in the expression analyses.

#### Multi-modal integration of NULISA proteomics and mass-cytometry cell counts

The following tuning strategy was performed for the NULISA inflammation panel. While pairwise correlations can be performed between proteomics-based analytes and mass cytometry-based cellular abundance, this does not fully capture the multivariate and coordinated response of the peripheral immune system. Therefore, we first performed a single Partial least squares (PLS) regression where NULISA features (X variables) were used to explain the abundance differences in cell counts (Y variables) (*Supplementary Figure 5A-B*). Based on the Q² error plot (*see below for Q^2^ calculation*), the first component was deemed sufficient [46] to construct the multiblock partial least squares (MB-PLS) model for integrating the two modalities (immune cell type counts and NULISA proteomics data).

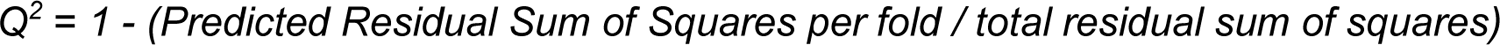

Several MB-PLS models (regression mode) were then constructed during optimization to determine number of variables required for the final model. A grid of the number of variables to select per modality was generated, and cross-modality correlations across paired components from each modality were measured. The number of variables retained was selected to maximize covariance of cross-modal components, while balancing the contributions of paired cross-modal components 1 and 2 (*Supplementary Figure 5A-B*). Cross-validation was performed using ‘M-fold’ (n=5 folds) with 50 repeats.

From the final integrated MB-PLS model, a bipartite network (i.e. edges connect variables from different modalities) was constructed, with |r|>= 0.4 as threshold. Diffusion propagation [47–49] was then performed to identify key plasma immune markers or peripheral cell types based on local network proximity. Heat diffusion propagation represents signal flow based on the local network topology. Diffusion time (extent of spread) *t*=0.1 was chosen to identify proximal variables, while maintaining strong localization signal from the inputs (where *t=0 represents no diffusion from inputs, and t=1 represents maximal diffusion*).

#### Linking immune cells with cognitive and clinical severity scores

We compared ACE-R cognitive scores between patients and controls (*Supplementary Figure 6*), and tested the association of scores for each ACE-R subdomain with disease duration (Years) in patients using loess smoothing (*Supplementary Figure 6*). Next, associations of immune cell type abundance and immune cluster scores with cognitive and clinical severity scores were evaluated. Partial correlation controlling for age, sex and Helios batches were computed with cell type abundance ratios as predictors (x) and each ACE-R subdomain or PSP-RS total scores as dependent variables (y). For each ACE-R subdomain score, p values of the analyses were FDR adjusted across all cell types. *Complex Heatmap* and *Circlize* R packages were used to show the resulting correlations in a heatmap.

#### Predictors of survival rates in patients

To identify key prognostic profiles of peripheral immune cells, a Ridge Cox model was constructed with cell types as predictors of survival (coded as: death = 1, alive at census = 0). Time intervals were calculated as days between date of blood sampling and date of death (if dead, n=8 patients) or date of census (if alive, n=52 patients; 4^th^ November 2025). Ridge regularization was applied equally for all cell types but not demographic covariates (e.g. age, sex) to prevent overfitting and to stabilize the estimates. The optimal lambda for the Ridge Cox model was selected via 5-fold cross validation. Bootstrapped sampling (1000 iterations) was performed to extract 95 percentile confidence intervals and p values.

As previous studies identified plasma NfL as robust proxy of disease progression and predictor of survival in patients with primary tauopathies [16, 25, 50–52], associations between individual peripheral immune cells and plasma NfL levels (NULISA NPQ units) were also tested in a regression model including all cell types as predictors and plasma NfL levels as outcome variable. A reduced regression model was identified via backward stepwise selection of the predictor variables, to reduce collinearity effect on convergence. Raw counts of cell type abundance were previously centered log ratio transformed against total live cells and then batch corrected using *removebatchEffect* function from *limma* R package.

## Results

### Cohort characteristics

No statistically significant group differences were observed in demographic variables (*See end of main text for Table 1, Supp Fig 3A-B*). No statistically significant differences were found in plasma brain-derived p-tau217 levels at group level between CBS, PSP, and control cohorts (*Kruskal Wallis across CBS, PSP, Control: p=0.14; Pairwise Dunn test PSP vs Control: FDR-corrected p=0.54, CBS vs Control: FDR-corrected p=0.15, PSP vs CBS: FDR-corrected p=0.13, Supp Fig 3A).* However, n=3 patients with CBS and n=1 patient with PSP, had high brain-derived p-tau 217 levels, identified as values > 2.5 standard deviations from the control mean (brain-derived p-tau 217: 1 SD_control_ =0.46; mean =10.8). No statistically significant differences were observed in plasma CRP levels between groups (*Two sided permutational t test, p=0.32, Supp Fig 3B*). Three of the patients had CRP levels between 2.5 and 3 standard deviations from the control mean (CRP: 1 SD_control_ = 1.08; mean =11.6).

**Table 1.** Cohort characteristics and group comparison statistics. For comparison between two groups: two-sided permutational tests (*t-test*), or Chi-square (categorial data). More than 2 groups comparison: Kruskal-Wallis and post-hoc Dunn test for pairwise comparisons.

|  | Control | PSP | CBS | Group Difference |
| --- | --- | --- | --- | --- |
| n | 21 | 60 |  | NA |
| Age, years, mean (SD) | 71 (6.06) | 73 (7.24) |  | *t-test*: t-statistic=1.07, p=0.29 |
| Sex (F/M) | 9/12 | 27/33 |  | Chi Square p=1.0, Chi square=9.87*10<sup>-31</sup> |
| Brain-derived p-Tau 217, mean (SD) | 10.8 (0.46) | 10.9 (0.6) | 11.7 (2.2) | Control vs PSP vs CBS:<br>Kruskal-Wallis's rank sum:<br>chi square = 3.93, p-value= 0.14<br>Dunn pairwise:<br>Control-CBS: Z = -1.95, FDR=0.15,<br>Control - PSP: Z= -0.60, FDR=0.54,<br>PSP-CBS: Z= -1.71, FDR=0.13 |
| C-reactive protein, mean (SD) | 11.6 (1.47) | 11.9 (1.62) |  | *t-test*: t-statistic=1.04, p=0.32 |
| Death by census date, count | 1 | 8 |  | NA |
| ACE-R Total score, mean (SD) | 96.6 (3.25) | 79.4 (15.5) |  | *t-test*: t-statistic= -7.21, p= 0.0001 |
| Attention and Orientation (ACE-R), mean (SD) | 18 (0) | 16.3 (2.73) |  | *t-test*: t-statistic= -4.36, FDR=0.0072 |
| Memory (ACE-R), mean (SD) | 25.2 (1.27) | 21.5 (4.92) |  | *t-test*: t-statistic= -4.64, FDR=0.00083 |
| Fluency (ACE-R), mean (SD) | 12.5 (2.02) | 6.39 (3.91) |  | *t-test*: t-statistic= -7.63, FDR=0.00050 |
| Language (ACE-R), mean (SD) | 25.6 (0.67) | 23.3 (3.29) |  | *t-test*: t-statistic= -4.66, FDR=0.0011 |
| Visuospatial (ACE-R), mean (SD) | 15.3 (1.07) | 11.8 (3.42) |  | *t-test*: t-statistic= -6.12, FDR=0.00050 |
| PSP-RS Total, median for PSP/CBS (SD) | NA | Median=36 (15.9) |  | NA |

Brain-derived p-Tau 217, C-reactive protein are in NULISA NPQ values. Unless stated otherwise, pairwise comparisons are patients (PSP/CBS) against controls as reference.

### Group differences in abundance of peripheral immune blood cells and cell type clusters

Group comparisons across all cell types found lower proportions (over total live cells) of Treg, Th17-like and Th2-like cells in patients as compared to controls (*Generalized negative binomial: Treg Group β = -0.59, FDR-corrected p=0.02, Th17-like Group β= -0.56, FDR-corrected p=0.02, Th2-like Group β= -0.70, FDR-corrected p=0.01, Fig 1A, Supp Table 1)*. Despite marked reductions in Treg and Th17-like cells, total CD4αβ+ T cell counts did not differ significantly between patients and controls. Lower proportions of transitional and nonclassical monocytes were found in patients, but similar abundance of classical monocytes was found between groups (*Generalized negative binomial: transitional monocytes Group β= -0.32, FDR-corrected p=0.014, nonclassical monocytes Group β= -0.48, FDR-corrected p=0.013, Fig 1A, Supp Table 1)*.

**Figure 1.**
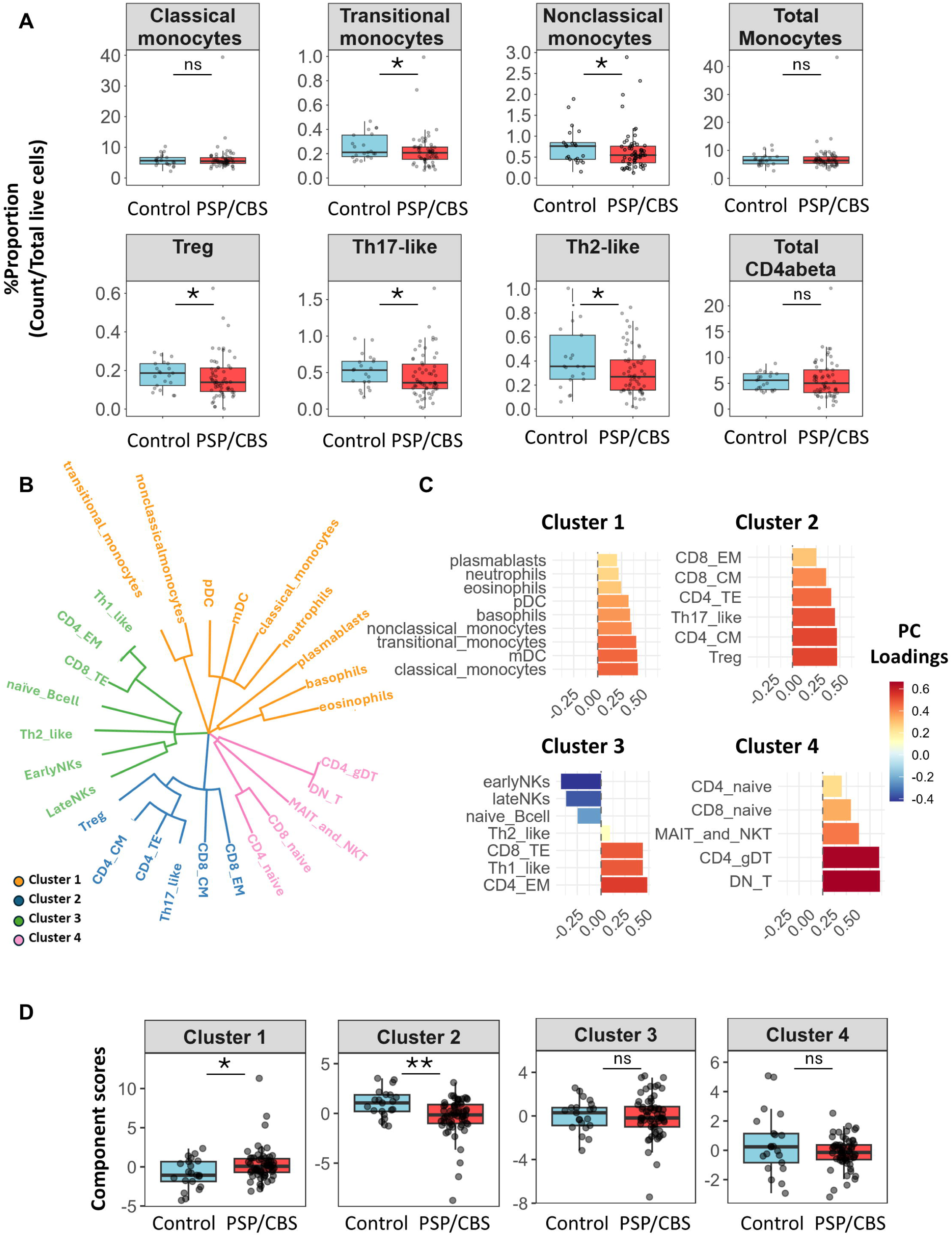
Differential abundance of peripheral immune cells in patients with PSP/CBS versus controls. (A) Proportions of peripheral immune cell populations in PSP/CBS and controls (cell-type count / total live cells per individual). *FDR < 0.05 in generalized negative binomial models adjusted for batch, sex, and age. (B) Hierarchical clustering dendrogram of immune cell types based on positive correlations in cell-type abundance. (C) Loadings of individual cell types on the first principal component within each cluster defined in (B). (D) Cluster’s component scores in PSP/CBS versus controls; each point represents one individual.

To further understand how the peripheral immune cells coordinate and covariate, we performed hierarchical clustering across cell types. The 27 cell types were divided into 4 separate clusters (*Fig 1B)* and the principal component analyses identified which cell types mostly contributed to each cluster *(Fig 1C*): Cluster 1 loaded onto classical, transitional and non-classical monocytes; Cluster 2 was represented by T αβ effector cells which influence autoimmunity such as Treg, Th17-like cells, CD4 Central Memory (CD4 CM); Cluster 3 grouped NK cells, effector T cells and naïve B cells, with CD8 Terminal Effector (CD8 TE), Th1-like and CD4 EM (CD4 Effector Memory) positively loading to the cluster; Cluster 4 included naïve CD4 and naïve CD8 cells, along with γδ T and DN T cells (*Fig 1B*). Patients with PSP/CBS had higher Cluster 1 scores (*Estimate=1.18, FDR-corrected p=0.02, Fig 1D*) and lower Cluster 2 scores (*Estimate= -1.31, FDR-corrected p=0.0016, Fig 1D*) than controls. No statistically significant group differences were found in cluster 3 and cluster 4.

### Increased covariance between immune cells in patients with PSP/CBS

We performed network analyses on correlations between cell types (count ratio as proportion of live cells), which can be used as proxy for immune cell coordination, in controls and patients, separately. Controls showed sparse connectivity between cells, while patients were characterized by a significant global increased covariance between immune cells *(Fig 2A)*. Directly comparing the networks across all cells between groups, patients with PSP/CBS showed a more than two-fold increase in number of significant (FDR<0.05, |r|>0.3) correlational links (*Wilcoxon rank sum: median_Controls_=1, median_PSP/CBS_ =5, FDR-corrected p<0.0001, Fig 2B, Supp Table 2*). Looking within covariance connectivity networks, clusters 1 and 2 were higher in patients, with cluster 2 showing the greatest change in number of significant correlations (>2.5fold higher in PSP/CBS group) (*Fig 2C, Supp Table 3, Wilcoxon rank sum: Cluster 1: median_Controls_=1.5, median_PSP/CBS_=3, FDR-corrected p=0.04, Cluster 2: median_Control_=2,median_PSP/CBS_=5.5, FDR-corrected p=0.007)*.

**Figure 2.**
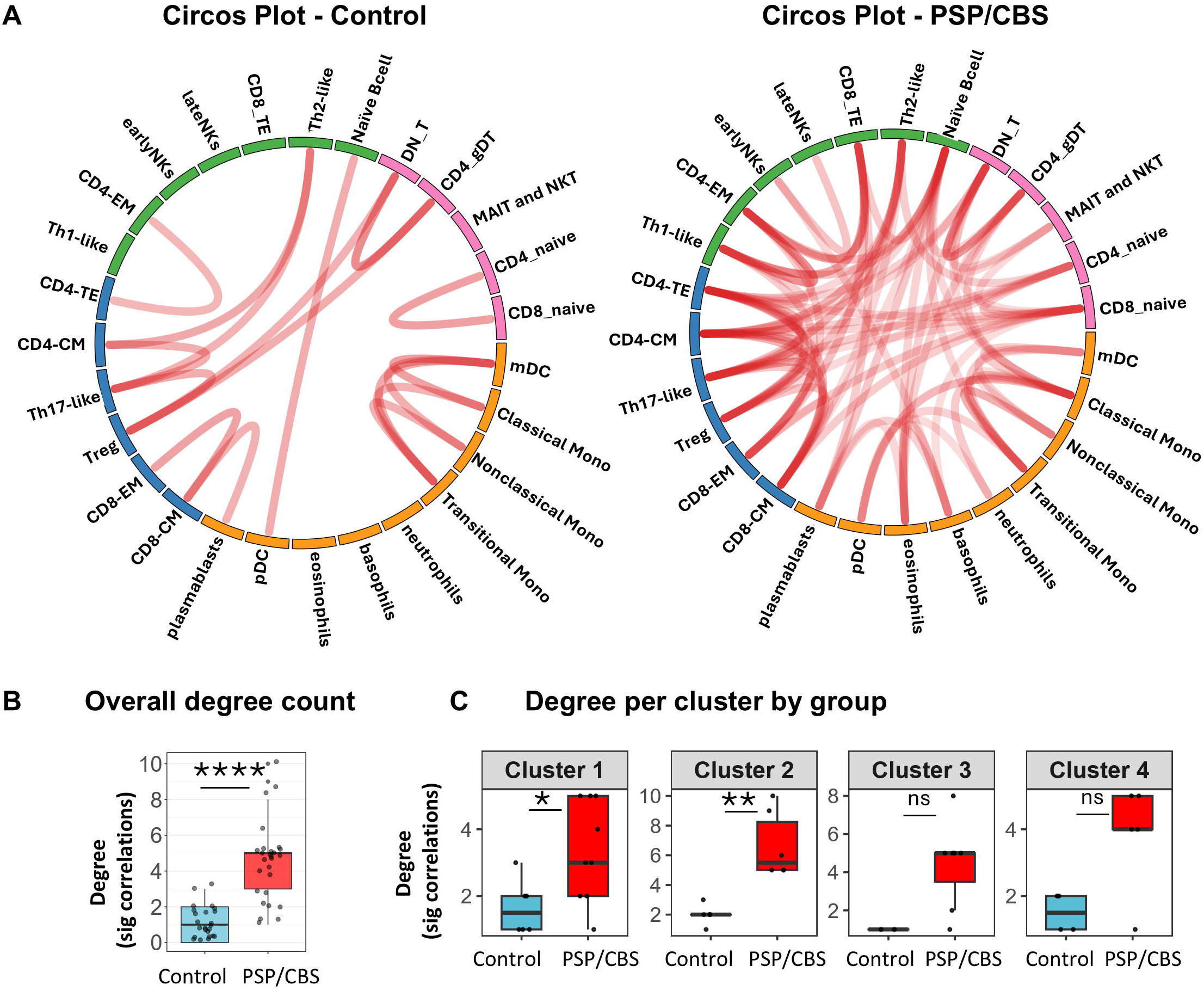
Increased covariance of immune cell-type networks in PSP/CBS. (A) Partial correlation networks of immune cell-type abundances in PSP/CBS and controls. Edges are partial Pearson correlations adjusted for age, sex, and batch, shown only if FDR < 0.05 and |r| > 0.3. (B) Total number of significant correlations (degree) in PSP/CBS versus control networks. (C) Degree per cell type within each cluster, by group (FDR < 0.05, |r| > 0.3).

Next, we employed metrics measuring the importance of each cell type in the network connectivity of controls or patients. Given the significant differences in total number of correlations between controls and patients, these metrics are normalized against the total degree of the respective networks. These metrics include (i) betweenness centrality, representing how often each cell type lies in the shortest paths of between other cell types within the network; and (ii) the weighted degree, the sum of absolute correlational coefficients (|r|) for correlations passing the threshold (FDR<0.05, |r|>0.3) (*Supp Fig 4*). In patients with PSP/CBS, the analysis revealed a drastic drop in betweenness centrality of Treg and Th17-like cells *(Supp Fig 4, Supp Table 4)*, as well as an increase in betweenness centrality of classical monocytes *(Supp Fig 4, Supp Table 4)*. This result was confirmed by bootstrapping with replacement, performed with 1000 iterations where partial correlations were recalculated per individual (*Supp Fig 4, Supp Table 4)*.

### Differential marker expression on monocytes, CD4 naïve and Th17 cells in patients with PSP/CBS

Within transitional monocytes, the patients showed higher expression of inflammation activation molecules such as CD197(CCR7) and CD161 [53, 54] (*Fig 3A-B, Supp Table 5*). No significant (FDR<0.05) differential expression was observed in classical or non-classical monocytes. In total monocytes, patients exhibited lower CD45RA and CD294 but higher CD197 and CD38 expression, in line with the loss of non-classical monocytes [55], which are typically antigen-presenting [55–57] (*Fig 3C, Supp Table 6*).

**Figure 3.**
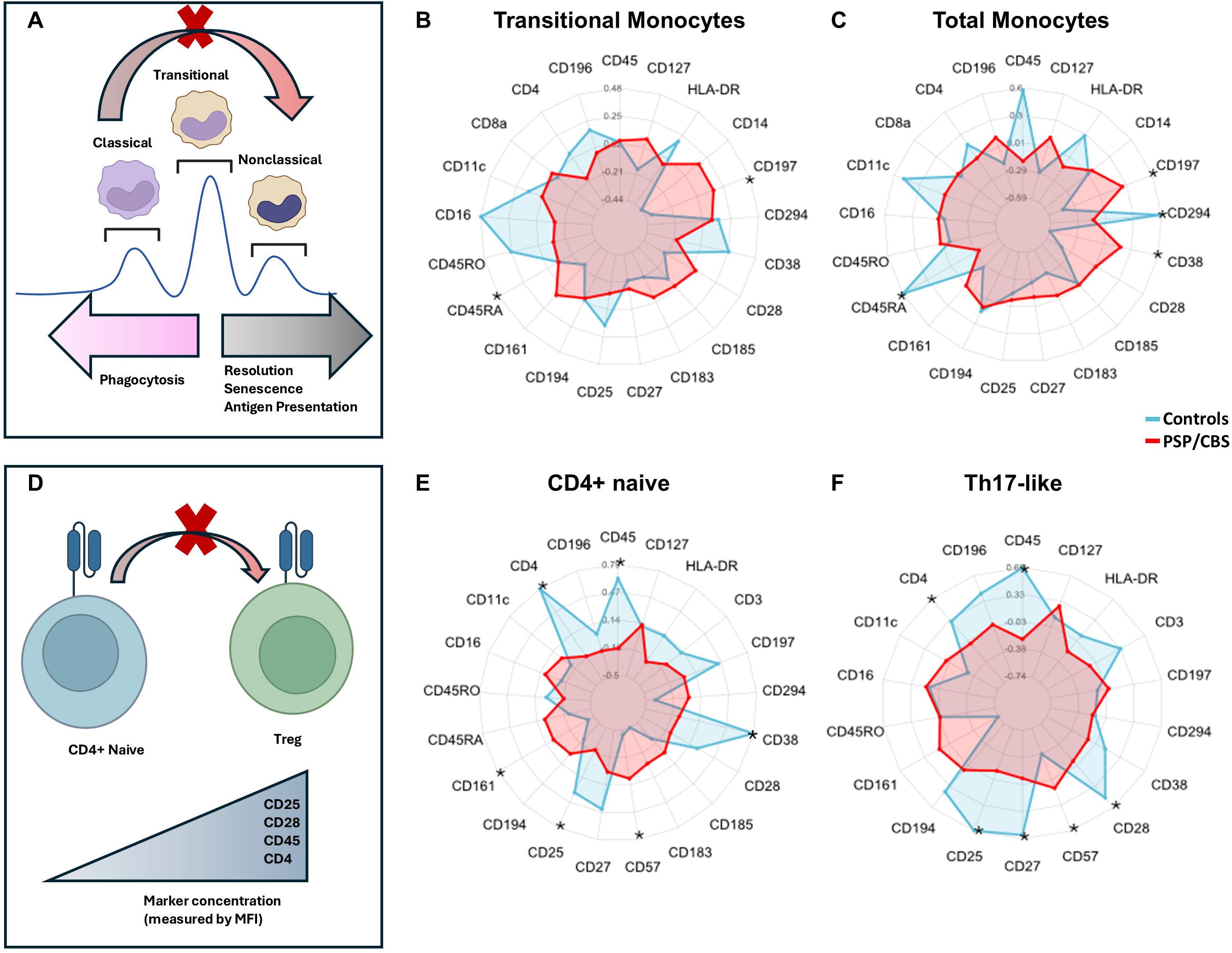
Dysregulated monocyte switching and impaired CD4–Treg differentiation in PSP/CBS. (A) Schematic of monocyte phenotypic switching from classical to non-classical via transitional monocytes, highlighting impaired transition in PSP/CBS. (B) Geometric median mass isotope intensity of markers in transitional and (C) total monocytes in PSP/CBS versus controls; *FDR-corrected p < 0.05. (D) Schematic of impaired CD4 T-cell differentiation to Treg in PSP/CBS, emphasizing expression of costimulatory and other signalling molecules required for differentiation to Treg. (E) Geometric median mass isotope intensity of markers in CD4 naïve T cells, and (F) Th17-like cells; *FDR-corrected p < 0.05.

Within the T cell populations, patients had reduced Treg counts but preserved CD4+ naïve cell numbers (*Fig 1A*). To understand if whether this may be driven by a dysfunction in Treg cell differentiation (from CD4 naïve cells), cell surface marker expression of CD4 naïve cells was compared between patients and controls. CD4 naïve cells in patients showed lower expression of CD45, CD4, CD38 and CD25, implying reduced availability of key costimulatory [58, 59] and activation signals [60, 61] required for Treg induction (*Fig 3D-E Supp Table 5*). In addition, Th17-like cells displayed lower CD27, CD25, CD45, CD28, CD4, but higher CD57 expression-pointing to a shift towards an exhausted pro-inflammatory effector state with impaired IL-17A–supporting co-stimulation [62–64] *(Fig 3D, Supp Table 5)*. No significant differential expression between the patients and controls were observed in Treg surface markers.

### Identification of key cytokines and immune molecules involved in intercellular communication between Treg, Th17-like and monocyte cell subpopulations

Given that Treg cells, Th17-like cells and monocyte cell subpopulations were reduced in patients with PSP/CBS, we investigated whether plasma inflammation markers, including cytokines, chemokines and immune regulation molecules, mediate the intercellular communication between these populations in patients. Thus, we included Treg, Th17-like and monocytic populations as starting inputs for the diffusion propagation analysis. Treg and Th17-like cells showed strong negative correlation with inflammation markers influencing autoimmune signalling such as IL-22, ANGPT1, IL-4, and GZMA/B, which are linked to efficacy of Treg cells controlling inflammation [65–71] (*Figure 4A*). In contrast, Treg and Th17-like cells showed strong positive correlations with CD276 (B7-H3), CCL21, IL-1R1 and GFAP, which are either involved immune tolerance or modulating Foxp3 activity in Treg cells (*Figure 4A*) [65–71].

**Figure 4.**
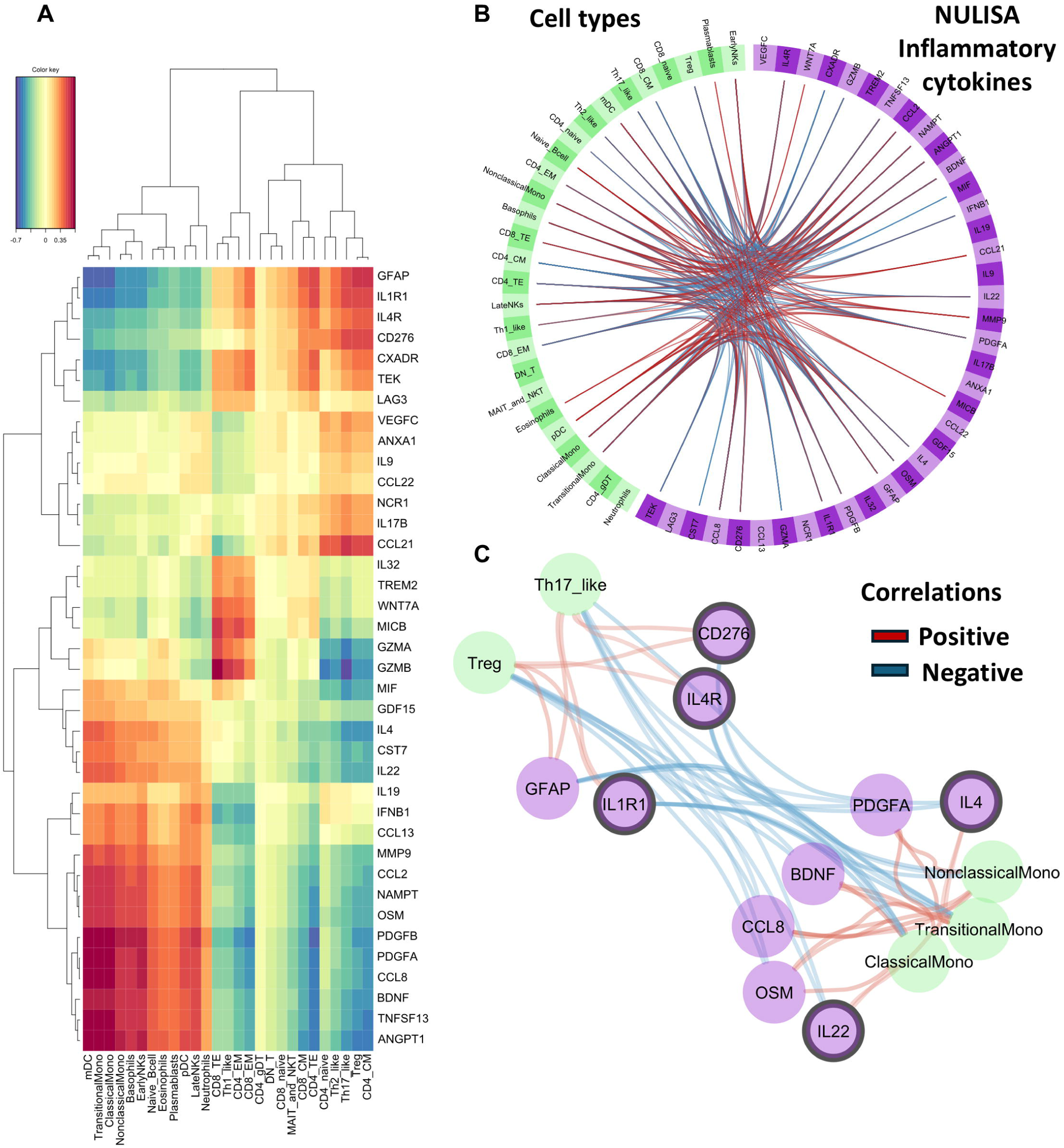
Plasma immune-regulatory molecules mediating intercellular communication between Treg/Th17-like cells and monocytic subpopulations. (A) Clustered image map of associations between NULISA inflammatory markers and immune cell-type counts from an MB-sPLS model; colour indicates association strength and direction. (B) Cross-modal network of cell types and plasma inflammation markers, thresholded at |r| ≥ 0.4 (blue, negative; red, positive). (C) Diffusion-based network prioritising cytokines and immune-modulatory molecules mediating signalling between Treg/Th17-like cells and monocytic subpopulations; Bold edges show prioritised links. Bordered nodes indicate the top five plasma immune-modulatory molecules.

We then constructed a cross-modal network from the integrated model, in which several immune cell types correlated with the immune-modulatory pattern of plasma markers (*Figure 4B*). Treg, Th17-like cells, along with classical and transitional monocytes were found to be the top 3 ranked hub cell types within the immune molecules-vs-cells integrated network as measured by network connectivity (*Supp Table 7*). Using Treg/Th17-like cells and monocytic subpopulations as diffusion inputs, we performed a diffusion propagation to prioritise key cytokines and immune molecules that likely mediate intercellular signalling between these populations (*Figure 4C, Supp Table 7*). These top ranked (n=5) immune-modulatory molecules and receptors include IL-22, IL-4, IL-4R, CD276, IL-1R1 (*Figure 4C, Supp Table 7; see Supplementary Figure 5C for group comparisons on these plasma markers*).

### Associations of peripheral immune cells with cognitive performance and clinical severity

To identify clinically relevant changes in peripheral immune cells, we tested for their association with ACE-R and PSP-RS scores (see Supp Fig 6 and Table 1 for group comparisons on the scores with controls). We found that Treg and Th17-like cell proportions (over total live cells) positively correlated with ACE-R total scores *(Treg: rho= 0.32, p=0.008, FDR-corrected p=0.075, Th17-like: rho= 0.34, p=0.005, FDR-corrected p=0.069, Fig 5A)*. In contrast, plasmablasts negatively correlated with ACE-R total scores *(plasmablasts: rho= -0.39, p=0.0013, FDR-corrected p=0.037, Fig 5A)*. Memory performance (ACE-R memory sub-score) was positively correlated with Th17-like and CD4 TE (CD4 Terminal Effector) cells *(Th17-like: rho= 0.35, p=0.013, FDR-corrected p=0.074, CD4 TE: 0.43, p=0.00042, FDR-corrected p=0.0076)* and negatively associated with classical monocytes, plasmablasts and mDC *(classical monocytes:rho= -0.36, p=0.0034, FDR-corrected p=0.03, plasmablasts: rho= -0.42, p=0.00056, FDR-corrected p=0.0076, mDC: rho = - 0.32, p=0.011, FDR-corrected p=0.07)*. Considering the interdependence of immune cells functions, we next evaluated the relationship between the four data-driven identified cell type clusters and cognitive performance. Cluster 2 values positively correlated with Fluency, Memory, Attention/Orientation and total ACE-R scores, while Cluster 1 values negatively correlated with Memory scores (*Cluster 2: ACER Total: rho=0.39, FDR-corrected p=0.0067, Fluency: rho=0.31, FDR-corrected p= 0.055, Memory: rho= 0.41, FDR-corrected p=0.003, Attention and Orientation: rho= 0.32, FDR-corrected p=0.043, Cluster 1: Memory: rho= -0.34, FDR-corrected p=0.01*, *Fig 5B*).

**Figure 5.**
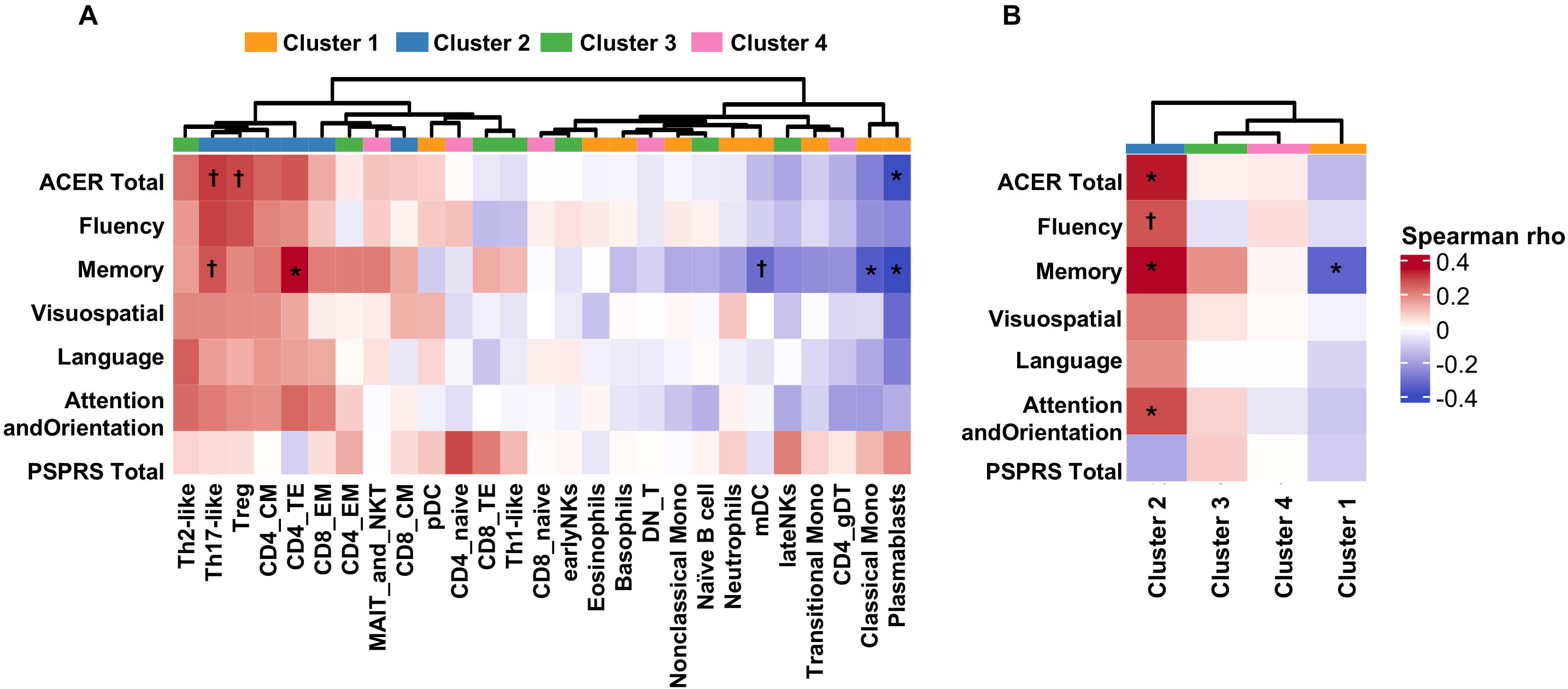
Associations between immune cell populations and clinical measures in PSP/CBS. (A) Heatmap of Spearman correlations between clinical capacity (ACE-R total and subdomains), disease severity (PSPRS), and immune cell-type abundances; *FDR-corrected p < 0.05. (B) Heatmap of correlations between clinical scores and cell-type cluster component scores; *FDR-corrected p < 0.05, †uncorrected p < 0.05.

### Peripheral immune cell alterations relate to plasma NfL and survival rates

We next examined associations between immune cell populations and disease progression, using plasma NfL as a marker of neurodegeneration associated with faster disease progression in PSP/CBS [16, 25, 50–52], and evaluating their predictive value for survival in patients. Using stepwise linear regression, higher frequencies of Th17-like cells and CD4 effector memory (CD4 EM), were associated with lower plasma NfL levels (*Fig 6A, Supp Table 8*). In contrast, higher CD4 terminal effector (CD4 TE) frequencies were associated with higher plasma NfL levels, consistent with an association between terminally differentiated CD4^+^ effector phenotypes and more advanced neurodegeneration (*Fig 6A, Supp Table 8*). Given that elevated plasma NfL predicts shorter survival in PSP and CBS, immune cell predictors of NfL were then evaluated as prognostic markers. In Ridge Cox proportional hazards models, higher Treg, Th17-like cells, CD4 central memory (CD4 CM), and naïve B cell counts were associated with a reduced hazard of death (HR < 1), whereas higher Th1-like frequencies were associated with an increased hazard (HR > 1), indicating a survival disadvantage in patients with a Th1-skewed CD4 response (*Treg: HR = 0.78, FDR-corrected p= 0.016; Th17-like: HR = 0.78, FDR-corrected p= 0.016; CD4 CM: HR = 0.82, FDR-corrected p= 0.020; naïve B cells: HR = 0.70, FDR-corrected p= 0.039; Th1-like: HR = 1.38, FDR-corrected p= 0.032; Fig 6B, Supp Fig 7, Supp Table 9*). Several NfL-associated cell types also contributed strongly to the first principal component of their respective immune clusters, prompting cluster-level analyses. Cluster 1 scores positively predicted plasma NfL levels, linking this innate/adaptive aggregate signature to greater neuroaxonal injury (*Cluster 1: β = 1.29, p= 0.047; Supp Fig 8, Supp Table 10*). Conversely, Cluster 2 scores independently predicted longer survival in models including plasma NfL and its interaction with cluster scores, suggesting that the Cluster 2 configuration confers resilience beyond the effect of NfL alone (*Cluster 2: HR = 0.52, p= 0.026; Fig 6C, Supp Table 11, Supp Fig 9*).

**Figure 6.**
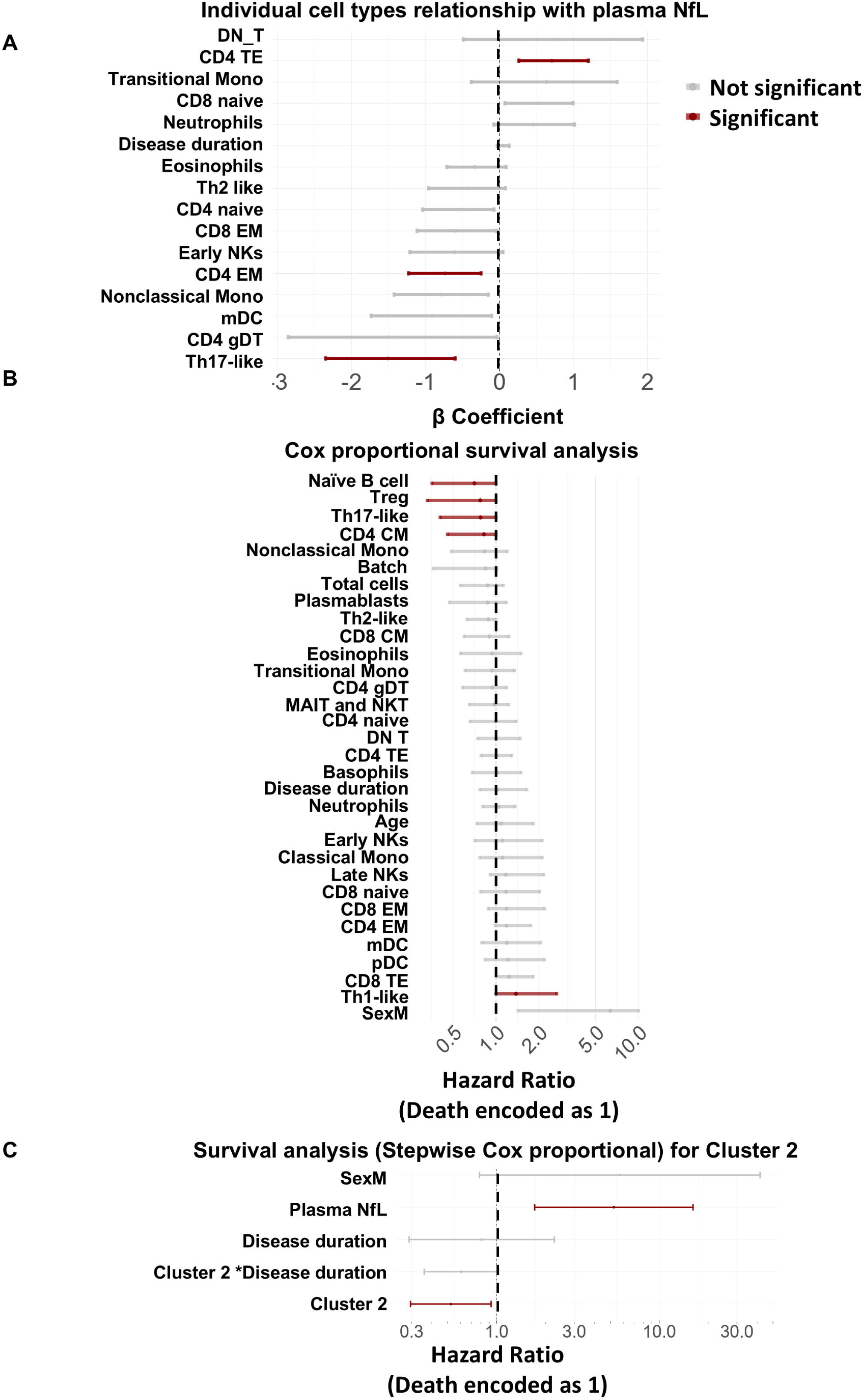
Peripheral immune alterations in relation to NfL and survival in PSP/CBS. (A) Stepwise linear regression of immune cell-type abundances against plasma NfL, adjusted for disease duration; significant predictors (FDR-corrected p < 0.05) are highlighted. (B) Ridge Cox models testing immune cell types as survival predictors; FDR-corrected p < 0.05 defines significance, with hazard ratios < 1 indicating lower death risk. (C) Stepwise Cox regression of Cluster 2 component scores and survival, including plasma NfL, disease duration, age, sex, and total cell counts; *p < 0.05.

## Discussion

Our findings demonstrate that individuals with PSP/CBS are characterised by dysregulated peripheral immune cell frequencies and network architectures that reflect impaired innate-adaptive immune crosstalk. Using high-dimensional mass cytometry and integrated multi-omics analysis, we identify two hallmark immune defects: (1) disrupted monocyte maturation indicated by the nonclassical and transitional monocytes depletion and (2) Treg depletion accompanied by profound loss of Treg network centrality in peripheral immune covariance structures. Network propagation analysis prioritized IL-22, IL-4, IL4R, IL-1R1 and CD276 as mediators of Treg-monocyte crosstalk, suggesting a dysregulated immune axis at the intersection of adaptive immune regulation and innate antigen presentation. These immunophenotypic alterations show associations with neurodegeneration (plasma NfL), cognitive decline, and mortality. These findings suggest that peripheral immune signatures may serve as fluid biomarkers for patient stratification and identify potential immunotherapy targets in primary tauopathies.

The immune system is highly functionally interconnected. As such, we implemented a system level and network learning approach to investigate peripheral blood immune signatures in patients with PSP/CBS. Cluster analyses identified two main immune profiles relevant in patients: an antigen-presenting monocytic cell cluster (Cluster 1) and an autoimmune regulation cluster (Cluster 2). *Cluster 1* grouped classical, transitional and nonclassical monocytes. Nonclassical monocytes are derived from classical monocytes via intermediary transitional phenotype [55, 57]. The higher betweenness centrality of classical monocytes the cell type covariance network of patients with PSP/CBS suggests that classical monocytes may serve as a hub in coordinating immune cells connectivity in these conditions. The shift towards exhausted centrally positioned classical monocytes may promote a chronically pro-inflammatory milieu, impair vascular surveillance normally mediated by patrolling nonclassical monocytes [72–74], and facilitate blood–brain barrier dysfunction and trafficking of pathogenic lymphocytes into selectively vulnerable midbrain regions [75–79]. Collectively, this pattern is compatible with a disruption of the canonical maturation continuum from classical to non-classical monocytes within patients with PSP/CBS. Expanding on earlier work [30, 80, 81], we show for the first time that classical monocytes are associated with worser cognitive functioning in individuals with PSP/CBS, suggesting that this maladaptive monocyte configuration is not only a biomarker of disease biology but also a potential contributor to disease progression.

*Cluster 2* captured marked reductions in Treg and Th17-like cells in patients with PSP/CBS, which was accompanied by a severe loss of connectivity within the Treg covariance network. This stands in stark contrast to cell covariance networks in healthy volunteers, where Treg cells function as key hub cell types coordinating peripheral immune homeostasis. The dual impairment - quantitative depletion and functional network isolation - suggests a fundamental breakdown in autoimmune regulation in PSP/CBS, with implications for both peripheral immune dysregulation and CNS pathology. Aligning with our findings, converging evidence supports a role for autoimmune mechanisms in PSP/CBS pathogenesis [77, 82]. Recent neuropathological studies reported striking infiltration of CD8+ cytotoxic T cells in the substantia nigra and midbrain tegmentum of PSP brains, with direct T cell-neuron contacts observed [77, 83]. Critically, CD8+ cell density did not correlate with disease duration or p-tau burden, suggesting this immune response represents a distinct pathophysiological feature rather than a secondary consequence of advanced neurodegeneration. Furthermore, tau epitopes were found binding to HLA in postmortem brain tissues of patients with PSP [82], and HLA haplotypes were shown to associate with distinct neuroinflammatory profiles and cytotoxic T cell densities, supporting an HLA-dependent autoimmune component to PSP pathogenesis [84]. Our findings of peripheral Treg depletion and loss of Treg network centrality expand the observations in CNS in earlier studies. In PSP/CBS, the combination of peripheral Treg depletion (reducing systemic autoimmune regulation) and CNS infiltration of cytotoxic CD8+ T cells may suggest a failure of peripheral autoimmune regulatory mechanisms that normally prevent tau-reactive T cells from becoming activated and trafficking to the brain [77, 82, 83, 85].

The parallel depletion of both Tregs and Th17-like cells in the PSP/CBS cohort warrants consideration. While Th17-like cells are traditionally viewed as pro-inflammatory and often positioned as functional antagonists of Treg cells [86–88], their relationship is more complex. Both effector cells can arise from CD4 naïve cells under TGF-β signalling, with IL-6 favouring differentiation to non-pathogenic Th17-like cells and IL-2 favouring Treg induction, and under chronic inflammatory conditions Tregs can convert to Th17-like cells (losing FoxP3, gaining RORγt), and vice versa, reflecting substantial plasticity between these lineages [89–91]. The coordinated depletion of both populations in PSP/CBS, together with downregulated CD25, CD45 and CD4 expression on CD4 naïve cells, therefore argues against a simple reciprocal balance and instead suggests a more fundamental defect in CD4⁺ T-cell differentiation and/or survival, potentially driven by chronic antigenic stimulation exhausting the naïve pool or by intrinsic impairment of co-stimulatory signalling required for both Treg and Th17-like cells induction. An alternative, non-exclusive possibility is that, under conditions of severely reduced Treg numbers, Th17-like cells may undergo compensatory phenotypic switching towards a regulatory phenotype in an attempt to restore immune homeostasis, a process canonically promoted by TGF-β and demonstrated in models where Th17 cells transdifferentiate into regulatory T cells during resolution of inflammation [90]. Future functional studies, including in vitro differentiation assays of isolated CD4 naïve cells and measurements of TGF-β production and plasticity in patient-derived Th17-like cells, will be essential to disentangle defective lineage commitment from compensatory lineage reprogramming in PSP/CBS.

Within the integrated cross-modality network, Treg, Th17-like cells, and classical and transitional monocytes were identified as hub cell types linked to immune-modulatory plasma molecules and other cell types in PSP/CBS. Linked to these cell populations, we identified a set of plasma inflammatory markers (including CD276, IL22, IL4, IL1R1) at the interface between Treg/Th17-like cells and monocytic populations, consistent with coordinated intercellular signalling. Immune-modulatory molecules and cytokines in this data-driven defined pattern have been previously shown to directly regulate Treg and Th17-like cell identity and function [65–71, 92, 93], and they exhibited the strongest correlations within the integrated network of cell type vs plasma markers in our cohort. Notably, several of these plasma markers showed opposite associations with Treg/Th17-like cells versus monocytes. For example, IL-1R1 levels were positively correlated with Treg abundance but negatively correlated with classical monocytes consistent with reports that higher IL-1R1 expression marks Treg with superior immunosuppressive function [65–71, 92, 93]. Likewise, IL4-R was positively associated with Treg cells and negatively associated with classical monocytes, in line with experimental evidence that IL-4/IL-4R signalling promotes an anti-inflammatory, nonclassical-like monocyte phenotype [94]. In the same module, the co-inhibitory checkpoint molecule CD276 (B7-H3) was also prominently represented, consistent with its emerging role as a context-dependent immune checkpoint at the T-cell–myeloid interface in neuroinflammatory and neurodegenerative settings [25, 95]. Together, these observations support a dysregulated peripheral Treg–monocyte axis in PSP/CBS, whereby altered cytokine and immune-regulatory signalling may link impaired regulatory T-cell function to maladaptive monocyte maturation and activation.

To explore the potential contributions of peripheral immune cells to neurodegeneration in PSP/CBS, we examined their associations with plasma NfL levels, a well-described marker of neuroaxonal injury and faster disease progression in PSP/CBS [16, 25, 50–52]. Th17-like cells showed the strongest negative association with plasma NfL. Non-pathogenic and host protective Th17-like cells have previously been described to express IL-10 and IL-17, whereas pro-inflammatory Th17-like preferentially express IL-22 together with IL-17 [96, 97]. In our dataset, Th17-like cells negatively correlated with IL-22, and showed concordant associations with several plasma immune-regulatory molecules that also tracked with Treg cells. These findings suggest that our analysis predominantly captures a non-pathogenic Th17-like phenotype, which may be linked to relative protection from neuroaxonal injury..

We further evaluated how immune cell alterations at the cluster level relate to clinical outcomes. Treg-driven Cluster 2 was positively associated with global cognition and conferred a survival advantage, remaining predictive of longer survival even after adjusting for NfL. In contrast, the monocyte-driven Cluster 1 was linked to poorer prognosis, showing a positive association with plasma NfL and aligning higher classical-dominated monocyte signatures with greater neuroaxonal injury. Together, these findings indicate that peripheral immune configurations, summarised by Cluster 1 and Cluster 2, capture biologically meaningful axes of vulnerability and resilience that map onto both molecular markers of neurodegeneration and clinical trajectories in PSP/CBS. In line with previous work using inflammation PET and cytokine composite measures [16, 18], composite scores of peripheral immune cells derived from Cluster 1 and Cluster 2 could complement NfL as stratification tools to identify patients at higher risk of rapid progression or to improve the sensitivity of future immunomodulatory trials in PSP/CBS.

Importantly, participants with systemic inflammation, autoimmune conditions, cancer, and recent vaccinations or infections were excluded from our study, and plasma C-reactive protein levels did not differ between patients and controls. Thus, the peripheral immune signatures we identify (including Treg/Th17-like cells depletion, disruption of Treg-driven and monocyte-driven clusters, and widespread rewiring of immune-cell covariance) are unlikely to reflect confounding systemic inflammatory stressors and instead appear intrinsic to PSP/CBS pathophysiology. This interpretation is further supported by their strong associations with disease-relevant outcomes: several immune features spanning both Cluster 1 and Cluster 2 predicted higher plasma NfL, worse cognitive performance, and shorter survival, and Cluster 2 scores independently predicted survival even after adjustment for NfL. Together, these observations suggest that peripheral immune dysregulation in PSP/CBS reflect pathogenic processes directly relevant to neurodegeneration rather than nonspecific by-products of systemic illness.

Our study has limitations. Firstly, diagnoses of PSP and CBS were based on clinical criteria rather than neuropathological confirmation, so a subset of cases may not be underpinned by primary tauopathies, but have alternative or mixed underlying pathologies that mimic PSP/CBS clinically. We attempted to mitigate effects related to non-neurodegenerative conditions by excluding individuals with pre-existing inflammatory, infectious, autoimmune, or malignant conditions that could independently alter immune regulation, and by screening for systemic inflammation, but some degree of diagnostic heterogeneity remains unavoidable in antemortem cohorts. Second, our molecular interpretations for specific cell populations are inferential and require orthogonal ex vivo validation in future work, for example functional characterization of patient-derived monocyte subsets and CD4⁺ populations (e.g. suppression assays, cytokine profiling, and differentiation studies). Third, although this is the largest mass-cytometry cohort in PSP/CBS, the sample size is modest for high-dimensional immune profiling and may increase variance in combinatorial analyses of parent and end-line populations, underscoring the need for replication in larger, independent cohorts to quantify the frequency and magnitude of autoimmune contributions to PSP/CBS. Finally, we were unable to directly assess IgLON5 or other CNS-restricted autoantibodies in brain tissue because all data were acquired antemortem; however, we sought to minimize inclusion of IgLON5-spectrum and other systemic autoimmune mimics by excluding patients with overt autoimmune disease and by confirming that plasma CRP levels did not differ between PSP/CBS and controls (*Supp Fig. 1*). Despite these limitations, our study provides one of the first high-dimensional characterizations of peripheral innate and adaptive immune cell alterations in PSP/CBS using human samples. Although the current data are insufficient to establish definitive causal mechanisms, they generate testable hypotheses and mechanistic models that can be pursued in targeted experimental systems, which is particularly important given that tauopathy-driven molecular processes cannot be fully recapitulated in existing animal models [98–100].

## Conclusion

In summary, we identified distinct peripheral blood immunophenotypic profiles in patients with PSP/CBS. Specifically, we provide novel evidence linking regulatory T cells, Th17-like cells, and disrupted monocyte phenotypic transitions to prognosis in PSP/CBS, with these immune populations associating closely with plasma neurodegeneration markers and survival. Patterns of correlation with plasma inflammatory mediators further suggest that Tregs exert functional effects that are often opposed to those of classical monocytes, consistent with a dysregulated Treg–monocyte axis. More broadly, our data reinforces the growing view that peripheral immune interactions may contribute directly to the pathogenesis of neurodegenerative diseases and, crucially, address a gap in understanding how peripheral immune perturbations translate into clinical outcomes in primary tauopathies. Collectively, our findings highlight key peripheral immune cell subsets which may serve as promising fluid biomarker candidates and potential immunomodulatory targets in PSP/CBS.

## Data Availability

All data produced in the present work are contained in the manuscript, and are available upon reasonable request to authors

## Acknowledgements

We thank our participant volunteers and their families for their participation in this study. We thank the National Institute for Health Research (NIHR) Cambridge BioResource centre staff, and the research nurses for their contribution, and the East Anglia Dementias and Neurodegenerative Diseases Research Network (DeNDRoN) for help with subject recruitment. We thank Cambridge BRC Cell Phenotyping Hub for their technical assistance in running the mass cytometry.

For the purpose of open access, the authors have applied a Creative Commons Attribution (CC BY) license to any Author Accepted Manuscript version arising from this submission. This work is licensed under a Creative Commons Attribution 4.0 International License.

## Author Contributions

Conception and funding: MM; design of the study: KOL, MM; methodology: KOL, MM, SLK, NSY; data acquisition: JG, HC, RF, SLK, NSY, PS, RD, JW, WL, TR, AH, MM; data analysis: KOL, MM; data interpretation: KOL, AH, JBR, HZ, JP, JTO, MM; writing - original draft: KOL, MM; writing - editing: KOL, JG, HC, RF, SLK, NSY, PS, RD, JW, WL, HP, AH, TR, JBR, HZ, JP, JTO, MM. All authors read and approved the final manuscript.

