## Supplementary Materials for "Peripheral Treg-monocyte immune signatures relate to neurodegeneration and prognosis in patients with primary tauopathies"

Cambridge CB2 0SZ, UK

**Supplementary Figure 1. Gating Hierarchy of broad peripheral immune cell types.** The gating strategy followed the one derived from *Maxpar Direct Immune Profiling* application protocol [43]. The icons were generated using Biorender.com.

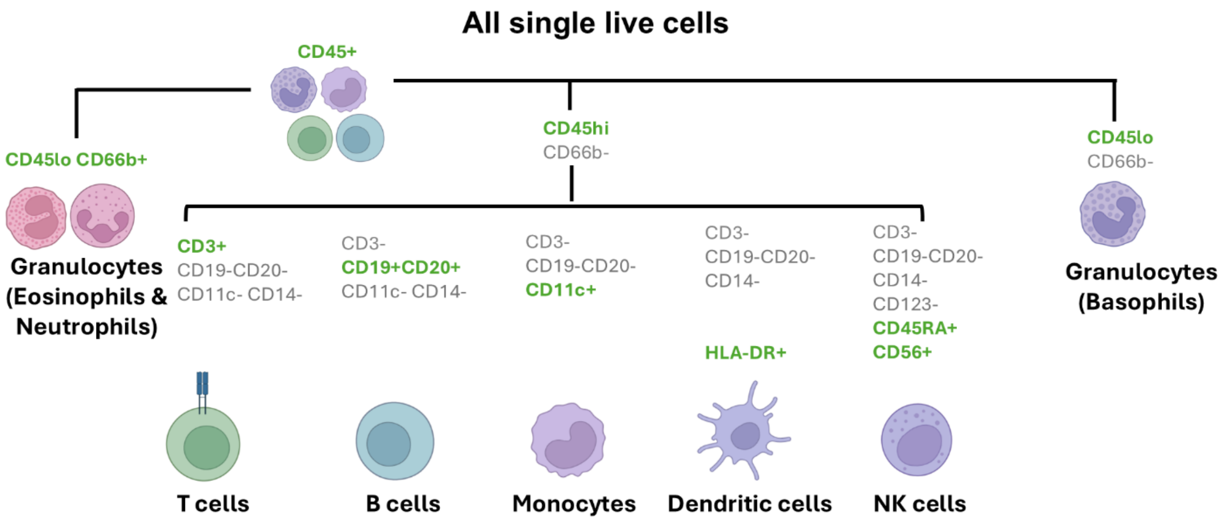

**Supplementary Figure 2. Gating Hierarchy of the peripheral immune cell types classified by lineages.** Labels in bold identify the individual cell types used for analyses. The gating strategy followed the one derived from *Maxpar Direct Immune Profiling* application protocol [43]. The icons were generated using Biorender.com.

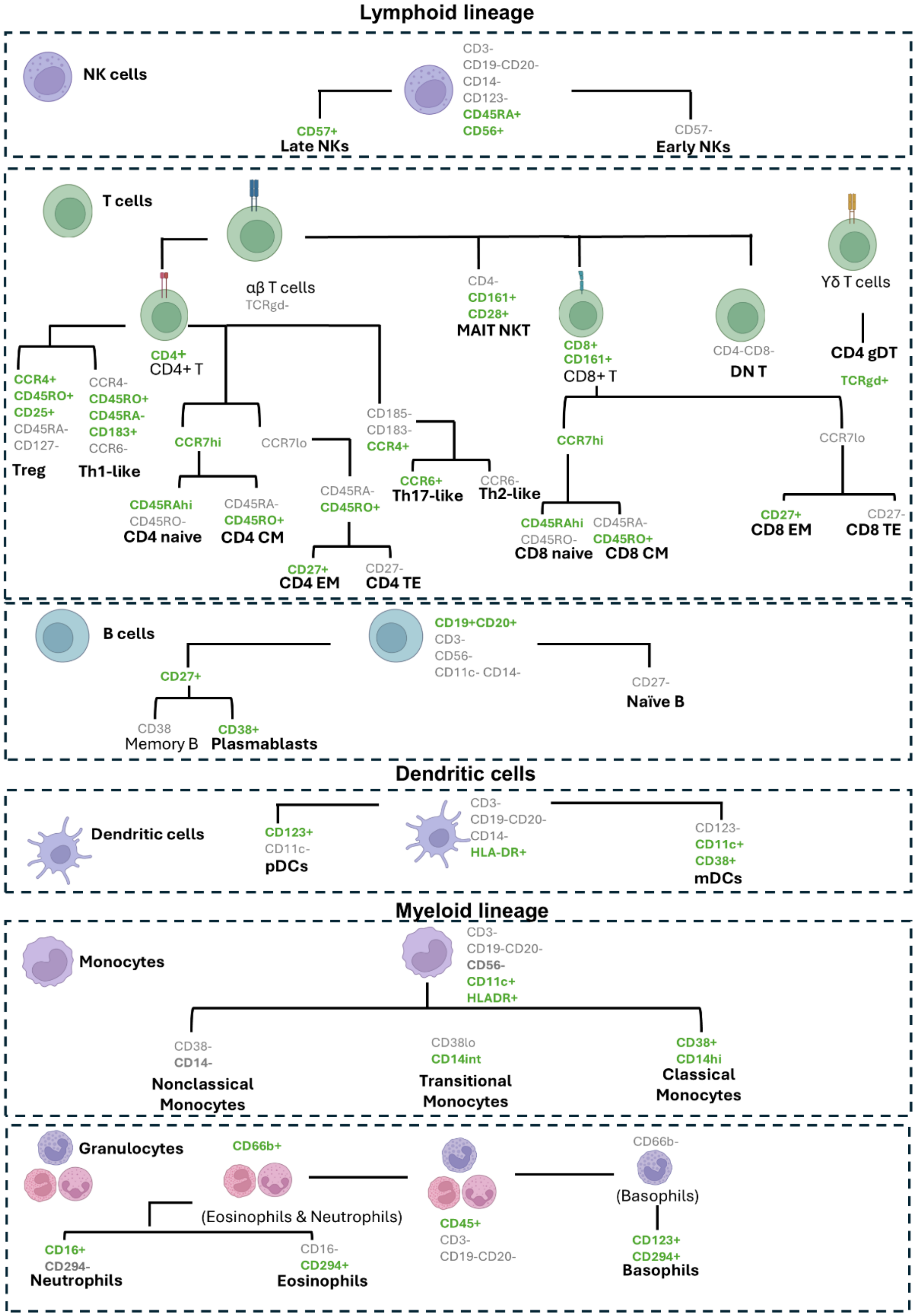

**Supplementary Figure 3. Demographics and plasma biomarker differences by groups.** (A) Brain-derived plasma pTau217 levels in controls, patients with CBS and PSP. Group differences were assessed with a Kruskal–Wallis's test ($p = 0.14$), followed by pairwise Dunn tests (PSP vs control: FDR-corrected $p = 0.54$; CBS vs control: FDR-corrected $p = 0.15$; PSP vs CBS: FDR-corrected $p = 0.13$). (B) Plasma CRP (normalised protein quantification) across groups; two-sided permutation t-test, $p = 0.32$.

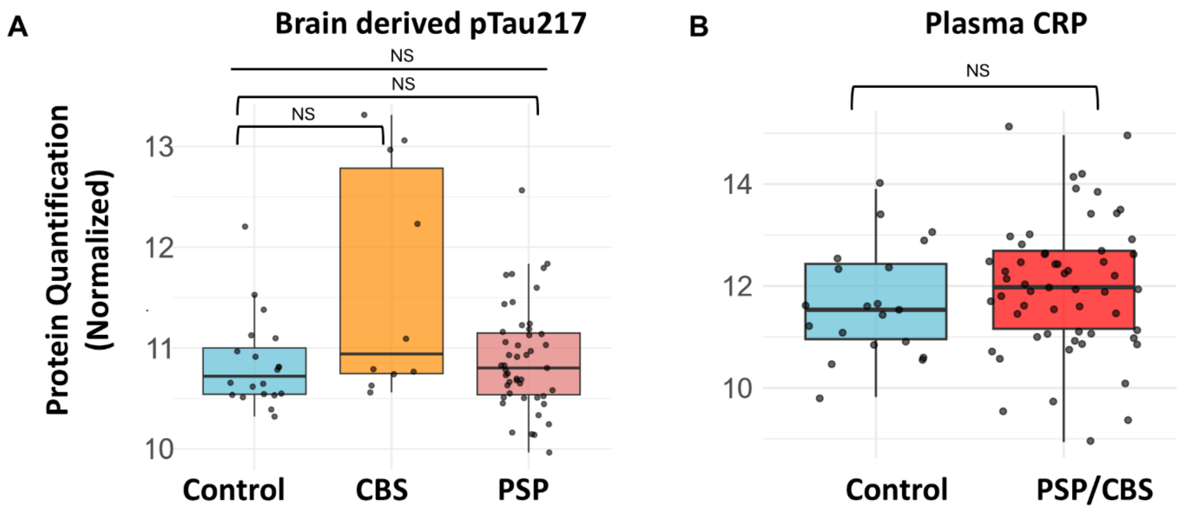

**Supplementary Figure 4. Hub cell types in PSP/CBS and control immune networks.** Hub cell types were defined as the top five nodes by betweenness centrality (higher values indicate greater network importance) within each group, measured on the cell-type correlation networks shown in Figure 2A. Bordered nodes indicate hub cell types. Betweenness centrality has been normalised with the number of significant correlations (FDR<0.05, |r|>0.3) in each respective network.

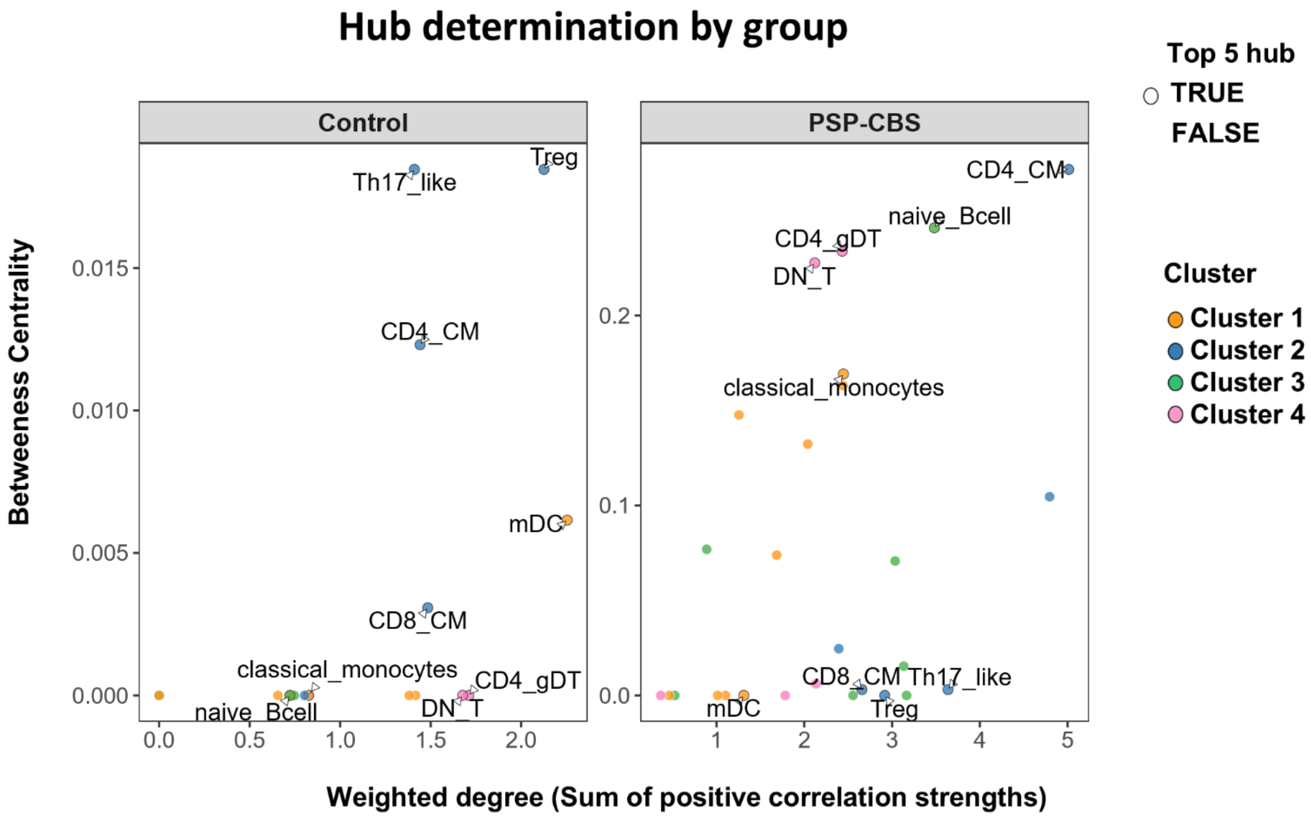

**Supplementary Figure 5. Tuning of integrated cross-modal models for construction of cytokine-cell type network**. (A) Tuning strategy for the NULISA inflammation panel to select the number of components in single sPLS; Q2 error plots are shown for models with NULISA cytokines as X and cell types as Y. Higher Q2 score refers to better predict capacity of the model where changes in inflammatory cytokines explain differences in cell type abundance. (B) Grid search for the number of variables retained per modality (cytokines vs cell types) based on cross modal correlations between paired components. (C) Protein abundance (normalized) for the top 5 cytokines and immune-modulatory molecules mediating signalling between Treg/Th17-like cells and monocytic subpopulations from Fig 4C. Protein abundance was tested for group differences using linear regression (Abundance ~ Group+Age+Sex+Batch), *** refers to p=0.00024.

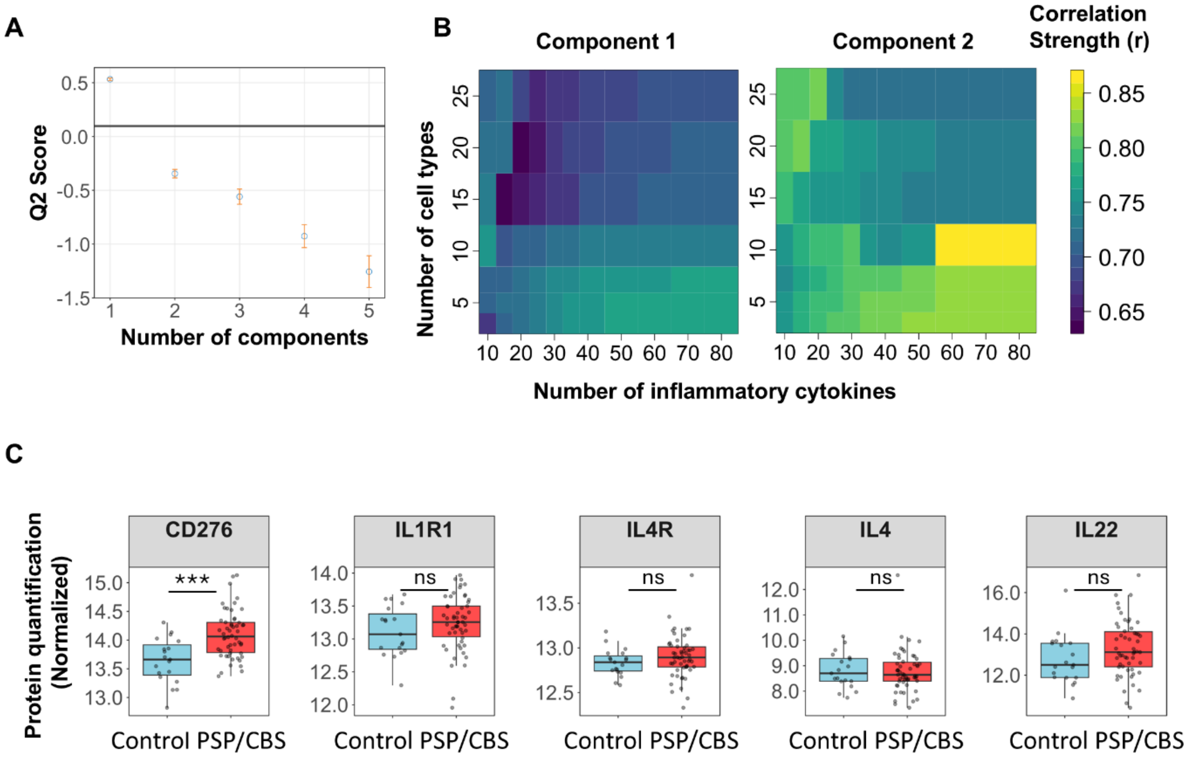

**Supplementary Figure 6. Cognitive profiles by groups.** (A) ACE-R total and subdomain scores in PSP/CBS versus controls. (B) Wilcoxon rank-sum tests comparing PSP/CBS and controls on ACE-R subdomain scores. (C) Nonlinear relationships between ACE-R subdomain scores and disease duration in PSP/CBS; r denotes Spearman's rho.

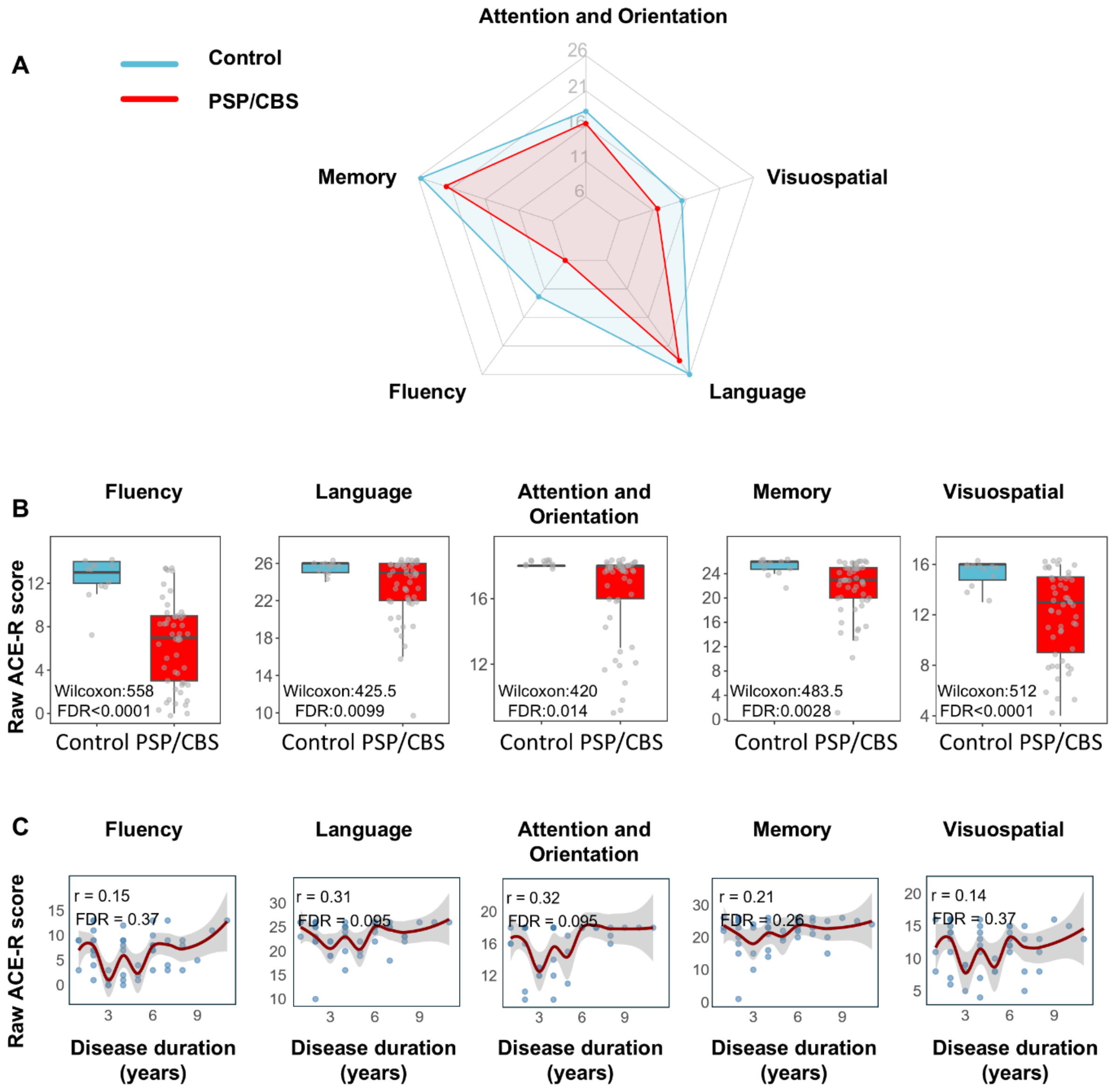

**Supplementary Figure 7. Kaplan-Meier survival curves for cell types previously identified to be significantly associated with survival in Fig 6B.** Patients were categorized into group with either high abundance or low abundance of the cell type. Abundance is split at the median of the respective cell type.

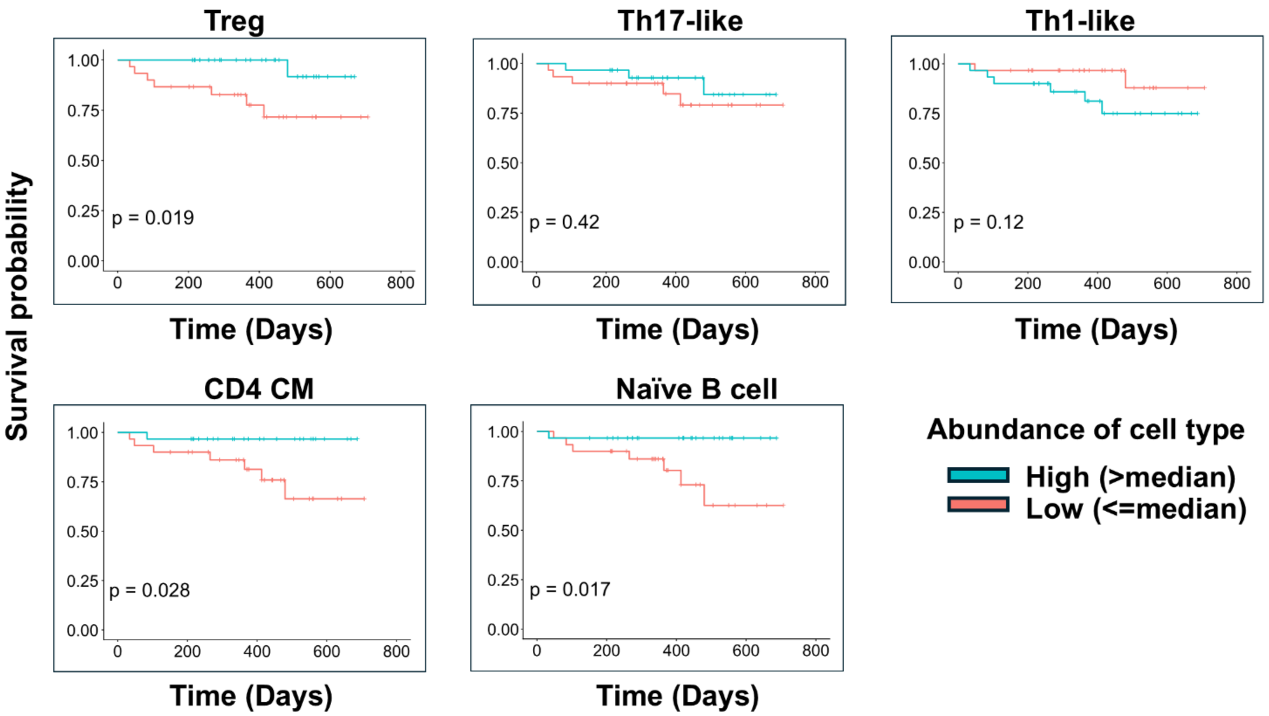

**Supplementary Figure 8. Immune cell clusters as predictors of plasma NfL.** Associations between immune cell cluster component scores and plasma NfL in PSP/CBS, modelled using a generalised linear model: plasma NfL ~ Cluster component scores * Age + Cluster component scores * Disease duration + Sex + Total cells.

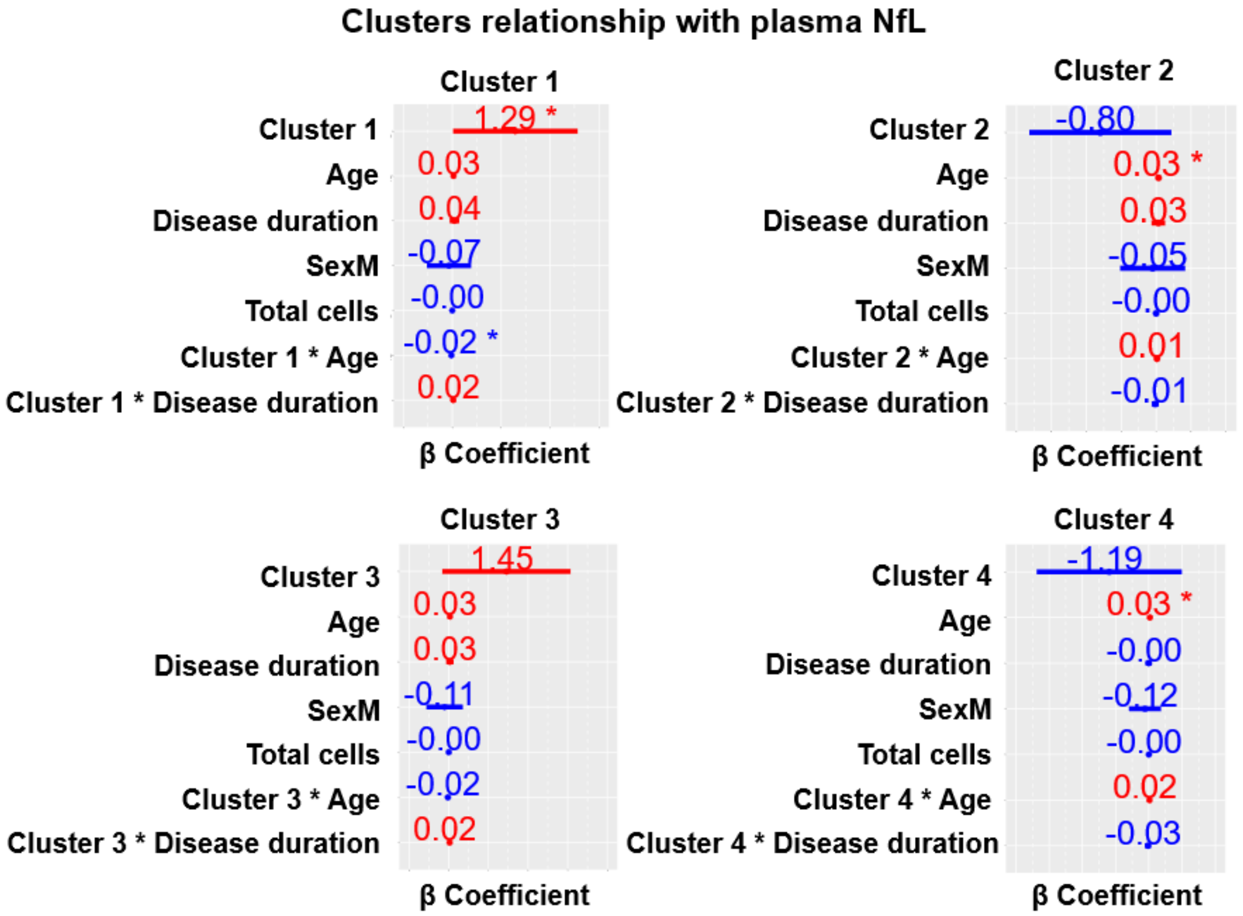

**Supplementary Figure 9. Stepwise covariate selection and full survival model**. Stepwise Cox modelling of survival including plasma NfL, disease duration, and their interactions with immune cluster component scores. Covariates were retained or removed based on change in AIC (ΔAIC), with negative ΔAIC indicating improved model fit.

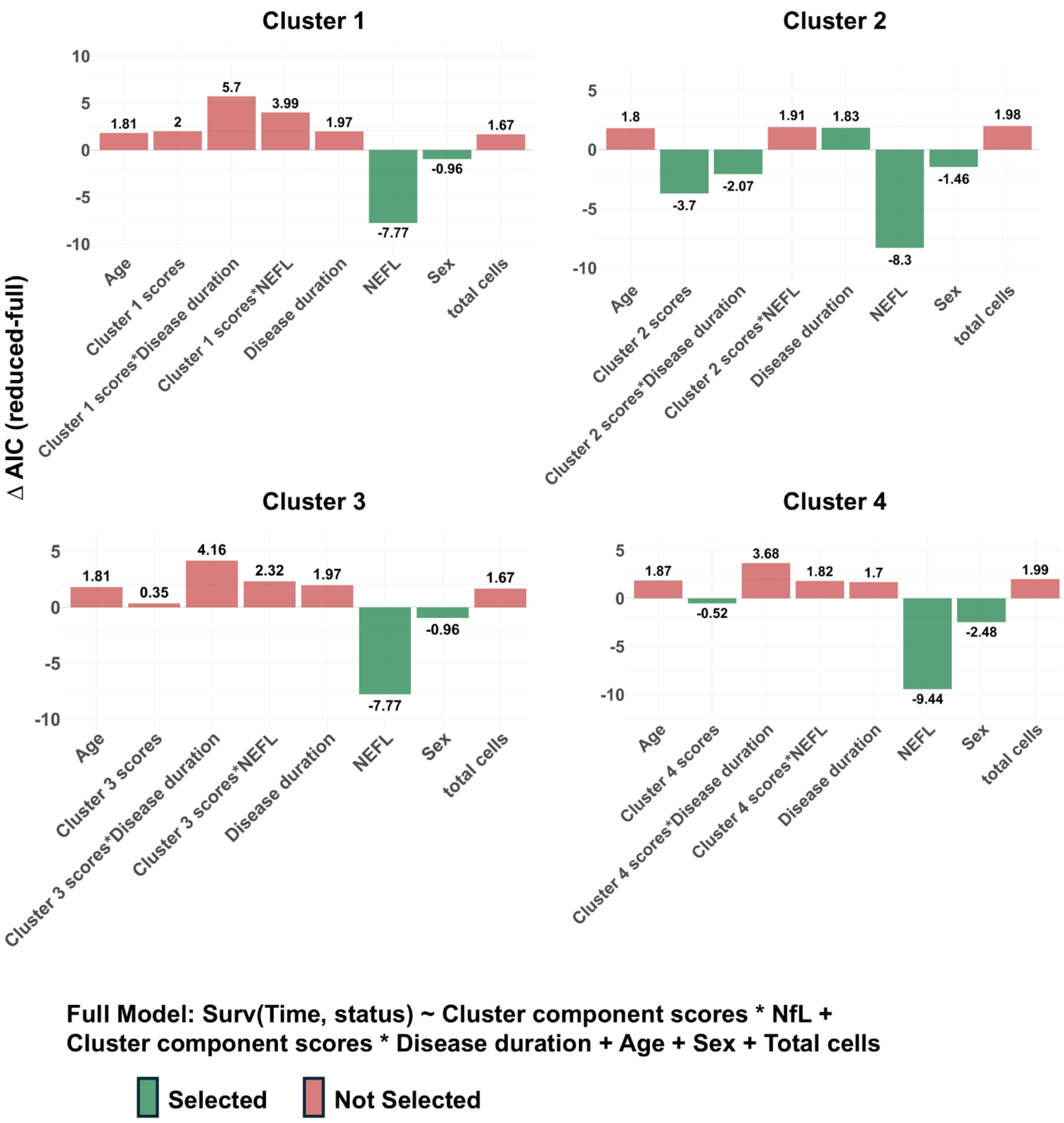

**Supplementary Table 1. Group comparisons on proportion of all peripheral immune blood cells (PSP/CBS vs Control).** Each point represents raw count of cell type/ total cells detected in that individual. Significance is based on generalized negative binomial model controlling for batch, sex, age. β-estimate reflects magnitude of coefficient for PSP/CBS vs Control.

| Cell type | β-estimate | p.value | FDR-corrected p |
|---|---|---|---|
| DN_T | -0.82 | 0.00015 | 0.0021 |
| CD4_gDT | -0.81 | 0.00016 | 0.0021 |
| Nonclassical Monocytes | -0.48 | 0.0017 | 0.013 |
| Th2_like | -0.70 | 0.0020 | 0.013 |
| Transitional Monocytes | -0.32 | 0.0026 | 0.014 |
| Treg | -0.59 | 0.0050 | 0.023 |
| Th17_like | -0.56 | 0.0070 | 0.027 |
| Early NKs | -0.32 | 0.036 | 0.11 |
| CD4_CM | -0.41 | 0.038 | 0.11 |
| CD4_naive | -0.46 | 0.047 | 0.12 |
| pDC | -0.24 | 0.051 | 0.12 |
| naive_Bcell | -0.33 | 0.056 | 0.13 |
| CD8_CM | -0.40 | 0.11 | 0.22 |
| Classical Monocytes | 0.11 | 0.12 | 0.23 |
| CD4_TE | -0.37 | 0.14 | 0.26 |
| mDC | -0.14 | 0.16 | 0.27 |
| MAIT/NKT | -0.36 | 0.19 | 0.29 |
| basophils | -0.17 | 0.20 | 0.29 |
| CD8_naive | -0.22 | 0.31 | 0.43 |
| CD8_TE | 0.35 | 0.33 | 0.44 |
| Th1_like | 0.28 | 0.34 | 0.44 |
| Late NKs | -0.19 | 0.39 | 0.47 |
| neutrophils | -0.081 | 0.43 | 0.51 |
| eosinophils | -0.15 | 0.46 | 0.51 |
| plasmablasts | -0.12 | 0.611 | 0.66 |
| CD4_EM | 0.14 | 0.66 | 0.68 |
| CD8_EM | -0.11 | 0.69 | 0.69 |

**Supplementary Table 2. Significant correlations between peripheral immune blood cells for each group (PSP/CBS and Controls).** Threshold for significance (|r|=>0.3, FDR-corrected p<0.05) for each individual peripheral immune blood cell types.

| Cell Type X | Cell Type Y | correlation | p_value | FDR | Group network |
|---|---|---|---|---|---|
| CD4_gDT | DN_T | 0.988 | 1.75E-14 | 6.14E-12 | Control |
| CD8_naive | CD4_naive | 0.827 | 2.28E-05 | 0.00282 | Control |
| Classical Monocytes | mDC | 0.826 | 2.41E-05 | 0.00282 | Control |
| CD8_EM | CD8_CM | 0.804 | 5.73E-05 | 0.00503 | Control |
| Th2_like | CD4_CM | 0.746 | 0.000377 | 0.0264 | Control |
| Transitional Monocytes | mDC | 0.731 | 0.000567 | 0.0311 | Control |
| CD4_gDT | Treg | 0.723 | 0.000698 | 0.0311 | Control |
| naive_Bcell | pDC | 0.722 | 0.000709 | 0.0311 | Control |
| Th17_like | Treg | 0.715 | 0.000862 | 0.0336 | Control |
| Nonclassical Monocytes | mDC | 0.697 | 0.00131 | 0.0431 | Control |
| Th17_like | CD4_CM | 0.696 | 0.00135 | 0.0431 | Control |
| DN_T | Treg | 0.688 | 0.0016 | 0.046 | Control |
| Transitional Monocytes | Nonclassical Monocytes | 0.685 | 0.0017 | 0.046 | Control |
| plasmablasts | CD8_CM | 0.68 | 0.00191 | 0.048 | Control |
| CD4_EM | Th1_like | 0.973 | 9.72E-37 | 3.41E-34 | PSP/CBS |
| CD4_gDT | DN_T | 0.918 | 1.07E-23 | 1.88E-21 | PSP/CBS |
| CD8_TE | CD4_EM | 0.814 | 1.3E-14 | 1.52E-12 | PSP/CBS |
| Th17_like | CD4_CM | 0.786 | 4.2E-13 | 3.68E-11 | PSP/CBS |
| CD8_TE | Th1_like | 0.77 | 2.64E-12 | 1.85E-10 | PSP/CBS |
| Th17_like | Treg | 0.744 | 3.38E-11 | 1.98E-09 | PSP/CBS |
| CD4_TE | CD4_CM | 0.719 | 2.89E-10 | 1.45E-08 | PSP/CBS |
| Transitional Monocytes | Nonclassical Monocytes | 0.698 | 1.56E-09 | 6.86E-08 | PSP/CBS |
| Th2_like | CD4_CM | 0.662 | 2.01E-08 | 7.83E-07 | PSP/CBS |
| CD8_CM | CD4_CM | 0.656 | 2.98E-08 | 1.05E-06 | PSP/CBS |
| CD4_CM | Treg | 0.646 | 5.66E-08 | 1.81E-06 | PSP/CBS |
| Classical Monocytes | pDC | 0.62 | 2.72E-07 | 7.96E-06 | PSP/CBS |
| Th17_like | Th2_like | 0.601 | 7.74E-07 | 2.09E-05 | PSP/CBS |
| Th17_like | CD4_TE | 0.564 | 4.83E-06 | 0.000121 | PSP/CBS |
| CD8_TE | CD8_EM | 0.554 | 7.96E-06 | 0.000186 | PSP/CBS |
| CD8_CM | CD4_TE | 0.549 | 9.61E-06 | 0.000211 | PSP/CBS |
| Transitional Monocytes | Classical Monocytes | 0.533 | 1.95E-05 | 0.000402 | PSP/CBS |
| naive_Bcell | Th1_like | 0.53 | 2.24E-05 | 0.000436 | PSP/CBS |
| naive_Bcell | CD4_EM | 0.528 | 2.44E-05 | 0.000451 | PSP/CBS |
| lateNKs | earlyNKs | 0.524 | 2.85E-05 | 0.000499 | PSP/CBS |
| CD8_CM | Treg | 0.513 | 4.55E-05 | 0.000761 | PSP/CBS |
| CD4_TE | Treg | 0.51 | 5.08E-05 | 0.000811 | PSP/CBS |

| | | | | | |
|---|---|---|---|---|---|
| Th2_like | Treg | 0.504 | 6.39E-05 | 0.000975 | PSP/CBS |
| CD8_EM | CD4_TE | 0.495 | 8.93E-05 | 0.00131 | PSP/CBS |
| basophils | eosinophils | 0.49 | 0.00011 | 0.0015 | PSP/CBS |
| plasmablasts | naive_Bcell | 0.49 | 0.000111 | 0.0015 | PSP/CBS |
| CD8_CM | Th17_like | 0.483 | 0.000141 | 0.00183 | PSP/CBS |
| CD8_naive | CD4_naive | 0.474 | 0.000193 | 0.00242 | PSP/CBS |
| Transitional Monocytes | mDC | 0.467 | 0.000252 | 0.00305 | PSP/CBS |
| Classical Monocytes | neutrophils | 0.461 | 0.000305 | 0.00348 | PSP/CBS |
| CD8_EM | CD8_naive | 0.46 | 0.000317 | 0.00348 | PSP/CBS |
| naive_Bcell | CD8_TE | 0.459 | 0.000327 | 0.00348 | PSP/CBS |
| Th17_like | CD4_naive | 0.459 | 0.000328 | 0.00348 | PSP/CBS |
| CD8_naive | CD8_CM | 0.456 | 0.000367 | 0.00379 | PSP/CBS |
| CD8_EM | CD4_EM | 0.446 | 0.000511 | 0.00512 | PSP/CBS |
| Classical Monocytes | mDC | 0.441 | 0.000598 | 0.00583 | PSP/CBS |
| CD8_EM | Th1_like | 0.439 | 0.00064 | 0.00603 | PSP/CBS |
| plasmablasts | CD8_TE | 0.438 | 0.000653 | 0.00603 | PSP/CBS |
| Th2_like | CD4_naive | 0.432 | 0.000793 | 0.00714 | PSP/CBS |
| CD4_TE | Th1_like | 0.422 | 0.00108 | 0.00949 | PSP/CBS |
| CD4_naive | CD4_CM | 0.418 | 0.00123 | 0.0105 | PSP/CBS |
| DN_T | eosinophils | 0.411 | 0.00151 | 0.0126 | PSP/CBS |
| DN_T | CD4_TE | 0.405 | 0.00177 | 0.0143 | PSP/CBS |
| CD4_TE | CD4_EM | 0.405 | 0.0018 | 0.0143 | PSP/CBS |
| Nonclassical Monocytes | mDC | 0.403 | 0.00187 | 0.0146 | PSP/CBS |
| plasmablasts | basophils | 0.403 | 0.00191 | 0.0146 | PSP/CBS |
| Transitional Monocytes | pDC | 0.394 | 0.00242 | 0.0181 | PSP/CBS |
| naive_Bcell | eosinophils | 0.392 | 0.00253 | 0.0185 | PSP/CBS |
| Classical Monocytes | CD4_gDT | 0.391 | 0.00266 | 0.0187 | PSP/CBS |
| CD4_gDT | eosinophils | 0.391 | 0.00266 | 0.0187 | PSP/CBS |
| DN_T | CD4_CM | 0.386 | 0.00304 | 0.021 | PSP/CBS |
| CD8_naive | CD4_CM | 0.374 | 0.00411 | 0.0278 | PSP/CBS |
| CD8_naive | CD4_TE | 0.368 | 0.00482 | 0.0313 | PSP/CBS |
| naive_Bcell | CD4_gDT | 0.368 | 0.00482 | 0.0313 | PSP/CBS |
| CD4_gDT | MAIT_and_NKT | 0.365 | 0.00526 | 0.0332 | PSP/CBS |
| naive_Bcell | CD4_CM | 0.365 | 0.00529 | 0.0332 | PSP/CBS |
| basophils | earlyNKs | 0.364 | 0.0054 | 0.0332 | PSP/CBS |
| plasmablasts | eosinophils | 0.356 | 0.00652 | 0.0392 | PSP/CBS |
| Th2_like | CD4_TE | 0.356 | 0.00659 | 0.0392 | PSP/CBS |
| naive_Bcell | Transitional Monocytes | 0.35 | 0.00769 | 0.045 | PSP/CBS |

**Supplementary Table 3. Number of significant correlations (degree) for each cell type for each group comparison and their respective cell type cluster** (previously defined in Fig 1B).

| Cell type | Degree (Control) | Degree (PSP/CBS) | Cell type Cluster |
|---|---|---|---|
| CD4_TE | 0 | 10 | 2 |
| CD4_CM | 2 | 9 | 2 |
| naive_Bcell | 1 | 8 | 3 |
| Eosinophils | 0 | 5 | 1 |
| Th1_like | 0 | 5 | 3 |
| CD4_EM | 0 | 5 | 3 |
| CD8_TE | 0 | 5 | 3 |
| Th2_like | 1 | 5 | 3 |
| Th17_like | 2 | 6 | 2 |
| CD8_naive | 1 | 5 | 4 |
| CD8_EM | 1 | 5 | 2 |
| Classical monocytes | 1 | 5 | 1 |
| CD4_naive | 1 | 4 | 4 |
| CD8_CM | 2 | 5 | 2 |
| CD4_gDT | 2 | 5 | 4 |
| basophils | 0 | 3 | 1 |
| Transitional monocytes | 2 | 5 | 1 |
| plasmablasts | 1 | 4 | 1 |
| Treg | 3 | 5 | 2 |
| DN_T | 2 | 4 | 4 |
| earlyNKs | 0 | 2 | 3 |
| Neutrophils | 0 | 1 | 1 |
| MAIT_and_NKT | 0 | 1 | 4 |
| pDC | 1 | 2 | 1 |
| lateNKs | 0 | 1 | 3 |
| mDC | 3 | 3 | 1 |
| Nonclassical monocytes | 2 | 2 | 1 |

**Supplementary Table 4. Group comparison of network metrics for each cell type.** Network metrics consist of sum of correlational strengths for significant correlation (weighted degree) and betweenness for each cell type. P_value and FDR represent calculation for bootstrapped betweenness comparing between the PSP/CBS and Control group. Significant threshold defined as FDR < 0.05 and |r| > 0.3). Cell type clusters are as previously defined in Fig 1B.

| Cell type | Cell type Cluster | Weighted degree (Control) | Weighted degree (PSP/CBS) | betweenness (Control) | betweenness (PSP/CBS) | betweenness (PSP/CBS-Control) | p_value | FDR |
|---|---|---|---|---|---|---|---|---|
| Th17_like | 2 | 1.41 | 3.64 | 0.019 | 0.0031 | -0.081 | <0.0001 | <0.0001 |
| Treg | 2 | 2.13 | 2.92 | 0.019 | 0 | -0.07 | <0.0001 | <0.0001 |
| pDC | 1 | 0.72 | 1.01 | 0 | 0 | -0.047 | <0.0001 | <0.0001 |
| CD4_naive | 4 | 0.83 | 1.78 | 0 | 0 | -0.04 | <0.0001 | <0.0001 |
| Th2_like | 3 | 0.746 | 2.56 | 0 | 0 | -0.034 | <0.0001 | <0.0001 |
| mDC | 1 | 2.25 | 1.31 | 0.0062 | 0 | -0.03 | <0.0001 | <0.0001 |
| CD4_EM | 3 | 0 | 3.17 | 0 | 0 | -0.028 | <0.0001 | <0.0001 |
| MAIT and NKT | 4 | 0 | 0.37 | 0 | 0 | -0.028 | <0.0001 | <0.0001 |
| Early NKs | 3 | 0 | 0.89 | 0 | 0.077 | -0.022 | <0.0001 | <0.0001 |
| Th1_like | 3 | 0 | 3.13 | 0 | 0.015 | -0.02 | <0.0001 | <0.0001 |
| Nonclassical monocytes | 1 | 1.38 | 1.1 | 0 | 0 | -0.014 | 0.0002 | 0.00025 |
| neutrophils | 1 | 0 | 0.46 | 0 | 0 | -0.0083 | 0.0018 | 0.0021 |
| CD8_CM | 2 | 1.48 | 2.66 | 0.0031 | 0.0031 | -0.0059 | 0.038 | 0.041 |
| CD8_EM | 2 | 0.8 | 2.39 | 0 | 0.025 | 0.0054 | 0.058 | 0.06 |
| CD8_naive | 4 | 0.83 | 2.13 | 0 | 0.0062 | 0.0062 | 0.094 | 0.094 |
| Late NKs | 3 | 0 | 0.52 | 0 | 0 | 0.0077 | 0.0024 | 0.0027 |
| CD8_TE | 3 | 0 | 3.03 | 0 | 0.071 | 0.011 | <0.0001 | <0.0001 |
| plasmablasts | 1 | 0.68 | 1.69 | 0 | 0.074 | 0.012 | <0.0001 | <0.0001 |
| Transitional monocytes | 1 | 1.42 | 2.44 | 0 | 0.16 | 0.028 | <0.0001 | <0.0001 |
| eosinophils | 1 | 0 | 2.04 | 0 | 0.13 | 0.031 | <0.0001 | <0.0001 |
| CD4_gDT | 4 | 1.71 | 2.43 | 0 | 0.23 | 0.036 | <0.0001 | <0.0001 |
| basophils | 1 | 0 | 1.26 | 0 | 0.15 | 0.049 | <0.0001 | <0.0001 |
| CD4_CM | 2 | 1.44 | 5.01 | 0.012 | 0.28 | 0.049 | <0.0001 | <0.0001 |
| CD4_TE | 2 | 0 | 4.79 | 0 | 0.11 | 0.062 | <0.0001 | <0.0001 |
| Classical monocytes | 1 | 0.83 | 2.45 | 0 | 0.17 | 0.065 | <0.0001 | <0.0001 |
| DN_T | 4 | 1.68 | 2.12 | 0 | 0.23 | 0.083 | <0.0001 | <0.0001 |
| naive_Bcell | 3 | 0.72 | 3.48 | 0 | 0.25 | 0.1 | <0.0001 | <0.0001 |

**Supplementary Table 5. Cell surface marker expression of transitional monocytes, Th17-like and CD4 naïve cells based on geometric median intensity** (FDR<0.05 threshold). Two-sided permutational testing was performed. T-statistic reflects group comparison (PSP/CBS – Control).

| Cell Type | marker | t_stat (PSP/CBS-Control) | p_value | FDR |
|---|---|---|---|---|
| CD4_naive | CD45 | -4.85 | 0.00000878 | 0.000184 |
| Th17_like | CD27 | -4.64 | 0.0000284 | 0.000512 |
| Th17_like | CD25_IL-2Ra | -4.3 | 0.000101 | 0.000911 |
| CD4_naive | CD57 | 4.16 | 0.0000948 | 0.000995 |
| Th17_like | CD45 | -3.89 | 0.000298 | 0.00167 |
| Th17_like | CD57 | 3.73 | 0.00037 | 0.00167 |
| CD4_naive | CD4 | -3.92 | 0.000289 | 0.00203 |
| CD4_naive | CD161 | 3.39 | 0.00112 | 0.005 |
| CD4_naive | CD38 | -3.48 | 0.00119 | 0.005 |
| CD4_naive | CD25_IL-2Ra | -3.01 | 0.00403 | 0.0141 |
| Transitional monocytes | CD161 | 3.37 | 0.00122 | 0.0157 |
| Transitional monocytes | CD197_CCR7 | 3.3 | 0.0015 | 0.0157 |
| Th17_like | CD28 | -3 | 0.00443 | 0.016 |
| Th17_like | CD4 | -2.7 | 0.00963 | 0.0289 |

**Supplementary Table 6. Cell surface marker expression for total monocyte based on geometric median.** Two-sided permutational testing was performed. T-statistic reflects group comparison (PSP/CBS – Control).

| Marker | t_stat (PSP/CBS-Control) | p_value | FDR |
|---|---|---|---|
| CD38 | 4.08 | 0.000243 | 0.0051 |
| CD45RA | -3.11 | 0.00383 | 0.0402 |
| CD294 | -2.76 | 0.00812 | 0.0479 |
| CD197_CCR7 | 2.71 | 0.00912 | 0.0479 |
| CD45 | -2.45 | 0.0191 | 0.0556 |
| CD11c | -2.49 | 0.0169 | 0.0556 |
| CD183_CXCR3 | 2.36 | 0.0212 | 0.0556 |
| HLA-DR | -2.49 | 0.0173 | 0.0556 |
| CD161 | 1.77 | 0.0823 | 0.192 |
| CD4 | -1.47 | 0.149 | 0.313 |
| CD28 | 1.28 | 0.207 | 0.383 |
| CD127_IL-7Ra | 1.25 | 0.219 | 0.383 |
| CD45RO | 1.05 | 0.303 | 0.49 |
| CD194_CCR4 | -0.458 | 0.65 | 0.91 |
| CD14 | 0.509 | 0.613 | 0.91 |
| CD196_CCR6 | 0.314 | 0.755 | 0.92 |
| CD8a | -0.124 | 0.902 | 0.92 |
| CD16 | -0.135 | 0.894 | 0.92 |
| CD25_IL-2Ra | 0.101 | 0.92 | 0.92 |
| CD27 | 0.321 | 0.751 | 0.92 |
| CD185_CXCR5 | 0.267 | 0.79 | 0.92 |

**Supplementary Table 7. Node properties of the cross modal integrated network between NULISA inflammation panel and cell types.** Nodes are either cell types or plasma inflammation markers from the NULISA inflammation panel. Network connectivity is ranked by degree and betweenness. Monocytic subpopulations, Treg and Th17-like are diffusion inputs (diffusion input=1) in the inflammation panel cross modal integrated network. Higher diffusion output refers to higher connectivity to the diffusion inputs. Diffusion output rank refers to node importance in connecting the diffusion inputs and are inclusive of diffusion inputs.

| Node in integrated network | data modality | diffusion input | diffusion output_heat | diffusion output_rank | Degree | Betweenness |
|---|---|---|---|---|---|---|
| Nonclassical monocytes | cell type | 1 | 0.308 | 0 | 15 | 0.0215 |
| Transitional monocytes | cell type | 1 | 0.26 | 1 | 18 | 0.0391 |
| Classical monocytes | cell type | 1 | 0.26 | 2 | 18 | 0.0391 |
| Th17_like | cell type | 1 | 0.251 | 3 | 18 | 0.0913 |
| Treg | cell type | 1 | 0.226 | 4 | 20 | 0.109 |
| IL4R | cytokine | 0 | 0.171 | 5 | 8 | 0.00611 |
| IL22 | cytokine | 0 | 0.148 | 6 | 6 | 0.00309 |
| CD276 | cytokine | 0 | 0.148 | 7 | 6 | 0.00309 |
| IL1R1 | cytokine | 0 | 0.145 | 8 | 13 | 0.0181 |
| IL4 | cytokine | 0 | 0.141 | 9 | 7 | 0.0102 |
| GFAP | cytokine | 0 | 0.139 | 10 | 14 | 0.0376 |
| OSM | cytokine | 0 | 0.136 | 11 | 15 | 0.0255 |
| BDNF | cytokine | 0 | 0.136 | 12 | 15 | 0.0255 |
| PDGFA | cytokine | 0 | 0.131 | 13 | 16 | 0.0522 |
| CCL8 | cytokine | 0 | 0.131 | 14 | 16 | 0.0522 |
| TNFSF13 | cytokine | 0 | 0.131 | 15 | 16 | 0.0376 |
| NAMPT | cytokine | 0 | 0.126 | 16 | 12 | 0.0145 |
| CCL2 | cytokine | 0 | 0.126 | 17 | 12 | 0.0145 |
| ANGPT1 | cytokine | 0 | 0.126 | 18 | 17 | 0.0566 |
| PDGFB | cytokine | 0 | 0.122 | 19 | 18 | 0.0904 |
| MMP9 | cytokine | 0 | 0.118 | 20 | 7 | 0.00184 |
| TEK | cytokine | 0 | 0.112 | 21 | 9 | 0.00472 |
| CXADR | cytokine | 0 | 0.112 | 22 | 9 | 0.00472 |
| mDC | cell type | 0 | 0.0942 | 23 | 18 | 0.0391 |
| CD4_CM | cell type | 0 | 0.0905 | 24 | 20 | 0.109 |
| MIF | cytokine | 0 | 0.0859 | 25 | 3 | 0.000176 |
| CST7 | cytokine | 0 | 0.0859 | 26 | 3 | 0.000176 |
| CD4_TE | cell type | 0 | 0.0852 | 27 | 15 | 0.0215 |
| Basophils | cell type | 0 | 0.0805 | 28 | 14 | 0.0178 |
| EarlyNKs | cell type | 0 | 0.0805 | 29 | 14 | 0.0178 |

| | | | | | | |
|---|---|---|---|---|---|---|
| CCL21 | cytokine | 0 | 0.0781 | 30 | 5 | 0.0143 |
| LateNKs | cell type | 0 | 0.0758 | 31 | 14 | 0.0392 |
| GZMA | cytokine | 0 | 0.0742 | 32 | 6 | 0.0325 |
| GZMB | cytokine | 0 | 0.0677 | 33 | 8 | 0.0723 |
| CD8_EM | cell type | 0 | 0.0671 | 34 | 12 | 0.0303 |
| Naive_Bcell | cell type | 0 | 0.0654 | 35 | 9 | 0.00468 |
| CD8_CM | cell type | 0 | 0.0654 | 36 | 9 | 0.00468 |
| pDC | cell type | 0 | 0.0631 | 37 | 9 | 0.00457 |
| Th2_like | cell type | 0 | 0.0295 | 38 | 5 | 0.00682 |
| CD4_EM | cell type | 0 | 0.0267 | 39 | 7 | 0.0768 |
| Eosinophils | cell type | 0 | 0.0263 | 40 | 3 | 0.000324 |
| Plasmablasts | cell type | 0 | 0.018 | 41 | 2 | 0.000102 |
| CD4_naive | cell type | 0 | 0.0135 | 42 | 3 | 0.00141 |
| CD8_TE | cell type | 0 | 0.00848 | 43 | 5 | 0.0181 |
| IFNB1 | cytokine | 0 | 0.00726 | 44 | 5 | 0.0273 |
| Th1_like | cell type | 0 | 0.00453 | 45 | 3 | 0.0044 |
| MICB | cytokine | 0 | 0.00157 | 46 | 3 | 0.00287 |
| WNT7A | cytokine | 0 | 0.00145 | 47 | 2 | 0.000893 |

**Supplementary Table 8. Stepwise regression evaluating relationship of peripheral immune blood cells in predicting plasma NfL levels.** β-estimate reflects magnitude of coefficient for change in NfL.

| Cell Type | β coefficient | 95%CI_Lower | 95%CI_Upper | p_value | FDR |
|---|---|---|---|---|---|
| Th17_like | -1.51 | -2.35 | -0.601 | 0.001 | 0.016 |
| CD4_TE | 0.706 | 0.261 | 1.2 | 0.003 | 0.024 |
| CD4_EM | -0.736 | -1.23 | -0.251 | 0.007 | 0.0373 |
| CD4_naive | -0.538 | -1.04 | -0.0738 | 0.023 | 0.062 |
| CD8_naive | 0.538 | 0.0719 | 0.995 | 0.031 | 0.062 |
| CD8_EM | -0.59 | -1.12 | -0.0414 | 0.031 | 0.062 |
| mDC | -0.911 | -1.73 | -0.103 | 0.026 | 0.062 |
| Nonclassical Monocytes | -0.789 | -1.43 | -0.152 | 0.017 | 0.062 |
| CD4_gDT | -1.5 | -2.86 | -0.0344 | 0.044 | 0.0782 |
| earlyNKs | -0.606 | -1.21 | 0.0473 | 0.059 | 0.0944 |
| Eosinophils | -0.332 | -0.711 | 0.0897 | 0.097 | 0.137 |
| Neutrophils | 0.456 | -0.0782 | 1.01 | 0.111 | 0.137 |
| Th2_like | -0.43 | -0.961 | 0.0778 | 0.106 | 0.137 |
| Disease duration | 0.0536 | -0.0305 | 0.129 | 0.185 | 0.211 |
| DN_T | 0.786 | -0.491 | 1.93 | 0.242 | 0.242 |
| Transitional Monocytes | 0.633 | -0.383 | 1.59 | 0.242 | 0.242 |

**Supplementary Table 9. Ridge Cox survival analyses evaluating relationship of peripheral immune blood cells in predicting survival.** p values are calculated by performing two-sided bootstrap sign test (1000 iterations).

| Cell Type | p_value | HR | 95%CI_Lower | 95%CI_Upper | FDR |
|---|---|---|---|---|---|
| Treg | 0.001 | 0.775 | 0.329 | 1 | 0.016 |
| Th17_like | 0.001 | 0.779 | 0.405 | 1 | 0.016 |
| CD4_CM | 0.00201 | 0.823 | 0.456 | 1 | 0.0214 |
| Th1_like | 0.00402 | 1.38 | 1 | 2.67 | 0.0321 |
| naive_Bcell | 0.00602 | 0.705 | 0.355 | 1 | 0.0386 |
| CD8_TE | 0.0241 | 1.23 | 1 | 1.82 | 0.11 |
| Sex_numeric | 0.0221 | 6.33 | 1.43 | 6761 | 0.11 |
| Th2_like | 0.0382 | 0.883 | 0.626 | 1 | 0.153 |
| batch_numeric | 0.0743 | 0.847 | 0.362 | 1.01 | 0.264 |
| CD4_EM | 0.0984 | 1.19 | 0.988 | 1.75 | 0.315 |
| CD8_EM | 0.243 | 1.18 | 0.882 | 2.19 | 0.69 |
| pDC | 0.323 | 1.21 | 0.839 | 2.19 | 0.69 |
| lateNKs | 0.301 | 1.17 | 0.912 | 2.16 | 0.69 |
| Nonclassical Monocytes | 0.309 | 0.83 | 0.486 | 1.2 | 0.69 |
| totalcells | 0.275 | 0.873 | 0.565 | 1.12 | 0.69 |
| CD8_CM | 0.39 | 0.899 | 0.597 | 1.23 | 0.72 |
| CD8_naive | 0.444 | 1.18 | 0.782 | 2.02 | 0.72 |
| mDC | 0.45 | 1.19 | 0.796 | 2.06 | 0.72 |
| Classical Monocytes | 0.44 | 1.11 | 0.771 | 2.11 | 0.72 |
| plasmablasts | 0.39 | 0.874 | 0.471 | 1.18 | 0.72 |
| CD4_gDT | 0.574 | 0.941 | 0.584 | 1.19 | 0.87 |
| earlyNKs | 0.598 | 1.11 | 0.711 | 2.11 | 0.87 |
| eosinophils | 0.739 | 0.938 | 0.564 | 1.5 | 0.876 |
| neutrophils | 0.683 | 1.05 | 0.809 | 1.36 | 0.876 |
| CD4_naive | 0.687 | 0.99 | 0.658 | 1.38 | 0.876 |
| Transitional Monocytes | 0.729 | 0.939 | 0.607 | 1.35 | 0.876 |
| Age_study_entry | 0.695 | 1.09 | 0.735 | 1.83 | 0.876 |
| DN_T | 0.787 | 0.997 | 0.745 | 1.48 | 0.9 |
| MAIT_and_NKT | 0.851 | 0.974 | 0.651 | 1.23 | 0.939 |
| CD4_TE | 0.892 | 1.01 | 0.785 | 1.29 | 0.947 |
| Disease_duration | 0.918 | 1.03 | 0.771 | 1.65 | 0.947 |
| basophils | 0.966 | 1.02 | 0.68 | 1.5 | 0.966 |

**Supplementary Table 10. Generalized linear model testing the association between peripheral immune cell cluster and plasma NfL levels in PSP/CBS.** Model as follows: plasma *NfL ~ Cluster component scores*Age + Cluster component scores*Disease_duration + Sex + totalcells*.

| Term | Estimate | pValue |
|---|---|---|
| ***Cluster 1*** | | |
| Cluster scores | 1.29 | 0.047 |
| Age | 0.03 | 0.12 |
| Disease duration | 0.04 | 0.37 |
| SexM | -0.067 | 0.77 |
| Total cells | <0.0001 | 0.91 |
| Cluster scores*Age | -0.02 | 0.042 |
| Custer scores*Disease duration | 0.02 | 0.40 |
| ***Cluster 2*** | | |
| Cluster scores | -0.8 | 0.12 |
| Age | 0.034 | 0.039 |
| Disease duration | 0.031 | 0.52 |
| SexM | -0.05 | 0.83 |
| Total cells | <0.0001 | 0.55 |
| Cluster scores*Age | 0.01 | 0.12 |
| Cluster scores*Disease duration | -0.01 | 0.59 |
| ***Cluster 3*** | | |
| Cluster scores | 1.45 | 0.078 |
| Age | 0.026 | 0.095 |
| Disease_duration | 0.026 | 0.57 |
| SexM | -0.11 | 0.65 |
| Total cells | <0.0001 | 0.94 |
| Cluster scores*Age | -0.02 | 0.056 |
| Cluster scores*Disease duration | 0.02 | 0.33 |
| ***Cluster 4*** | | |
| Cluster scores | -1.19 | 0.28 |
| Age | 0.03 | 0.045 |
| Disease_duration | -0.005 | 0.92 |
| SexM | -0.12 | 0.63 |
| Total cells | <0.0001 | 0.58 |
| Cluster scores*Age | 0.018 | 0.26 |
| Cluster scores*Disease duration | -0.028 | 0.55 |

**Supplementary Table 11. Stepwise Cox survival analysis evaluating the associations between peripheral immune cell cluster and survival in PSP/CBS.** Full model as illustrated in Supp Fig 9. HR<0 reflects terms which predict survival, while HR >0 reflects terms which predicts death.

| Term | HR | p_value |
|---|---|---|
| ***Cluster 1*** | | |
| NfL | 4.93 | 0.0028 |
| SexM | 3.82 | 0.11 |
| ***Cluster 2*** | | |
| Cluster scores | 0.52 | 0.026 |
| NfL | 5.28 | 0.0036 |
| Disease duration | 0.81 | 0.69 |
| SexM | 5.73 | 0.085 |
| Cluster scores*Disease duration | 0.61 | 0.063 |
| ***Cluster 3*** | | |
| NfL | 4.93 | 0.0028 |
| SexM | 3.82 | 0.11 |
| ***Cluster 4*** | | |
| Cluster scores | 1.83 | 0.87 |
| NfL | 6.95 | 0.0028 |
| SexM | 5.60 | 0.052 |
